# Standards for Reporting Diagnostic Accuracy Studies Using Intraoperative Neurophysiological Monitoring

**DOI:** 10.1101/2025.07.25.25331837

**Authors:** Parthasarathy D. Thirumala, Jeffrey R. Balzer, Andrea Szelenyi, Kathleen Seidel, Aatif M. Husain, Patrick M. Bossuyt, Gea Drost, the STARD-IONM Working Group

## Abstract

**Objective:** Intraoperative neurophysiological monitoring (IONM) plays a critical role in preserving neural function during surgery. Comparing IONM research studies is challenging due to methodological heterogeneity and inconsistent reporting. The Standards for Reporting Diagnostic Accuracy Studies Using Intraoperative Neurophysiological Monitoring (STARD-IONM) checklist was developed to improve transparency, reproducibility and comparability.

**Methods:** We used a four-phase consensus process. Phase 1 convened an expert steering group to define the rationale, plan the protocol, and establish the working group. Phase 2 adapted the STARD 2015 checklist to IONM, with feedback from studies. Phase 3 invited input from professional societies, and the broader community. Phase 4 focuses on dissemination and implementation.

**Results:** A review of 12 IONM studies using the 30-item STARD-2015 checklist revealed frequent underreporting of methods, missing data handling, adverse events, and blinding of index-test interpretation and outcome assessment. Based on these findings and expert consensus, we developed the STARD-IONM checklist. The checklist retains the STARD-2015 framework while adding IONM-specific reporting elements, including alert thresholds; signal-event classification as no deterioration, partial recovery, reversible deterioration, or irreversible deterioration; anesthesia and physiology reporting; and an evaluation framework to support standardized reporting. Dissemination and implementation are currently underway.

**Conclusions:** STARD-IONM extends STARD-2015 by providing specific reporting recommendations that address the methodological challenges in IONM diagnostic-accuracy research.

**Significance:** Adoption of STARD-IONM can enhance reporting quality, transparency, reproducibility, thereby facilitating more rigorous evaluation and comparison of IONM studies across surgical domains.

## Introduction

Intraoperative neurophysiological monitoring (IONM) encompasses a set of diagnostic techniques used to assess the functional integrity of the nervous system during surgeries that pose risk to neural function. It includes continuous monitoring modalities such as electroencephalography (EEG), electromyography (EMG), and evoked potentials (EPs), as well as mapping techniques to identify and protect eloquent cortical, subcortical, brainstem, spinal cord or peripheral nervous system structures.(Deletis et al., 2020; MacDonald et al., 2019, 2013) Significant intraoperative changes in these recordings may signal impending injury and guide real-time surgical interventions to reduce neurological deficits.(Nuwer et al., 1995; Radtke et al., 1989; Skinner and Sala, 2017; Skinner and Holdefer, 2014) IONM therefore serves as a valuable tool for enhancing the safety and outcomes of surgical procedures.(Sala et al., 2006; Skinner et al., 2017) In addition, intraoperative data can guide immediate postoperative management and may help to distinguish transient from permanent deficits. (Neuloh et al., 2004; Seidel et al., 2025; Szelényi et al., 2006)

Despite its widespread use, concerns remain regarding the diagnostic accuracy of IONM. Key limitations include variable outcome definitions, small sample sizes, unclear documentation of surgical responses to IONM changes, and methodological heterogeneity.(Asimakidou et al., 2021; Holdefer and Skinner, 2020; Nwachuku et al., 2015; Sala et al., 2007; See et al., 2016) Diagnostic accuracy refers to the agreement between changes detected by the IONM technique (the index test) and perioperative neurological outcomes (the reference standard).(Bossuyt et al., 2003) Although many studies report higher rates of neurological deficit among patients with significant or persistent IONM changes, variability in study design, thresholds, and reporting limits evidence synthesis and translation to practice.(Asimakidou et al., 2021) Systematic reviews, cohort studies, and guidelines have addressed some of these challenges and evaluated the diagnostic accuracy of EEG, somatosensory evoked potentials (SSEP), motor evoked potentials (MEPs), and brainstem auditory evoked potentials (BAEPs).(Chang et al., 2020; Dulfer et al., 2024; MacDonald et al., 2013; Neuloh et al., 2004; Seidel et al., 2025, 2013; Szelényi et al., 2006; Thirumala et al., 2014; Toleikis et al., 2024) However, conclusions are often constrained by methodological heterogeneity and limitations in reporting. While individualized intraoperative baselines are standard practice, there is no consensus on thresholds for clinically significant changes, which vary across institutions and modalities.(Chang et al., 2020; Dulfer et al., 2023) Furthermore, details regarding the timing and nature of surgical interventions in response to IONM changes are rarely reported,(Holdefer and Skinner, 2020) and perioperative outcome assessments lack uniformity, complicating evaluation of IONM’s diagnostic utility.

Several factors affect both the internal and external validity of IONM diagnostic accuracy studies. Internal validity refers to whether IONM changes truly reflect underlying neurological dysfunction. For example, systematic reviews of SSEPs and MEPs in detecting cerebral ischemia often do not specify when intraoperative baselines are established,(Reddy et al., 2018) which is important because factors such as anesthesia induction, hemodynamic parameters, and patient demographics (e.g., age) can significantly influence EP amplitudes at the start of surgery.(Dulfer et al., 2025, 2024, 2023; Kawaguchi et al., 2020) External validity relates to the generalizability of study findings. In brain tumor surgery, for instance, mapping and monitoring aims to preserve eloquent cortex and tracts, but variability in techniques and limited reporting on tumor grade, cortical plasticity, or patient characteristics constrain broader applicability.(Gerritsen et al., 2026; Neuloh et al., 2007; Seidel et al., 2022; Szelényi et al., 2010; De Witt Hamer et al., 2012) This limitation is particularly relevant for preservation of language and other higher cognitive functions, as mapping approaches during awake surgery have evolved from focal localization models to hodotopical network-based frameworks.(Duffau, 2015; Sanai et al., 2008)

Given the complexity and variability of IONM, standardized reporting is essential. The STARD (Standards for Reporting of Diagnostic Accuracy) initiative, launched in 2003 and updated in 2015, provides a checklist and flow diagram to enhance transparency and reduce bias in diagnostic accuracy studies. (Bossuyt et al., 2003, 2015; Whiting et al., 2004, 2011) However, IONM poses unique challenges that warrant tailored guidance. Despite improvements in reporting following STARD,(Bossuyt et al., 2015, 2003; Cohen et al., 2016) its adoption in IONM literature has been limited. This gap is consequential because IONM studies are often interventional (signals can trigger intraoperative changes in management) and are influenced by anesthesia and physiological confounders. To address these gaps, the STARD-IONM initiative aims to improve the quality, transparency, and consistency of diagnostic accuracy studies in IONM by adapting STARD-2015 principles to the IONM context.(Bossuyt et al., 2015) By promoting comprehensive and standardized reporting, STARD-IONM seeks to reduce bias and enhance the applicability and generalizability of findings across clinical settings.

## Methods

We adapted the STARD framework to STARD-IONM following established guidance (STARD-2015, STARDdem) using a prespecified four-phase process (Figure 1).(Bossuyt et al., 2015; Noel-Storr et al., 2014)

**Figure 1.**
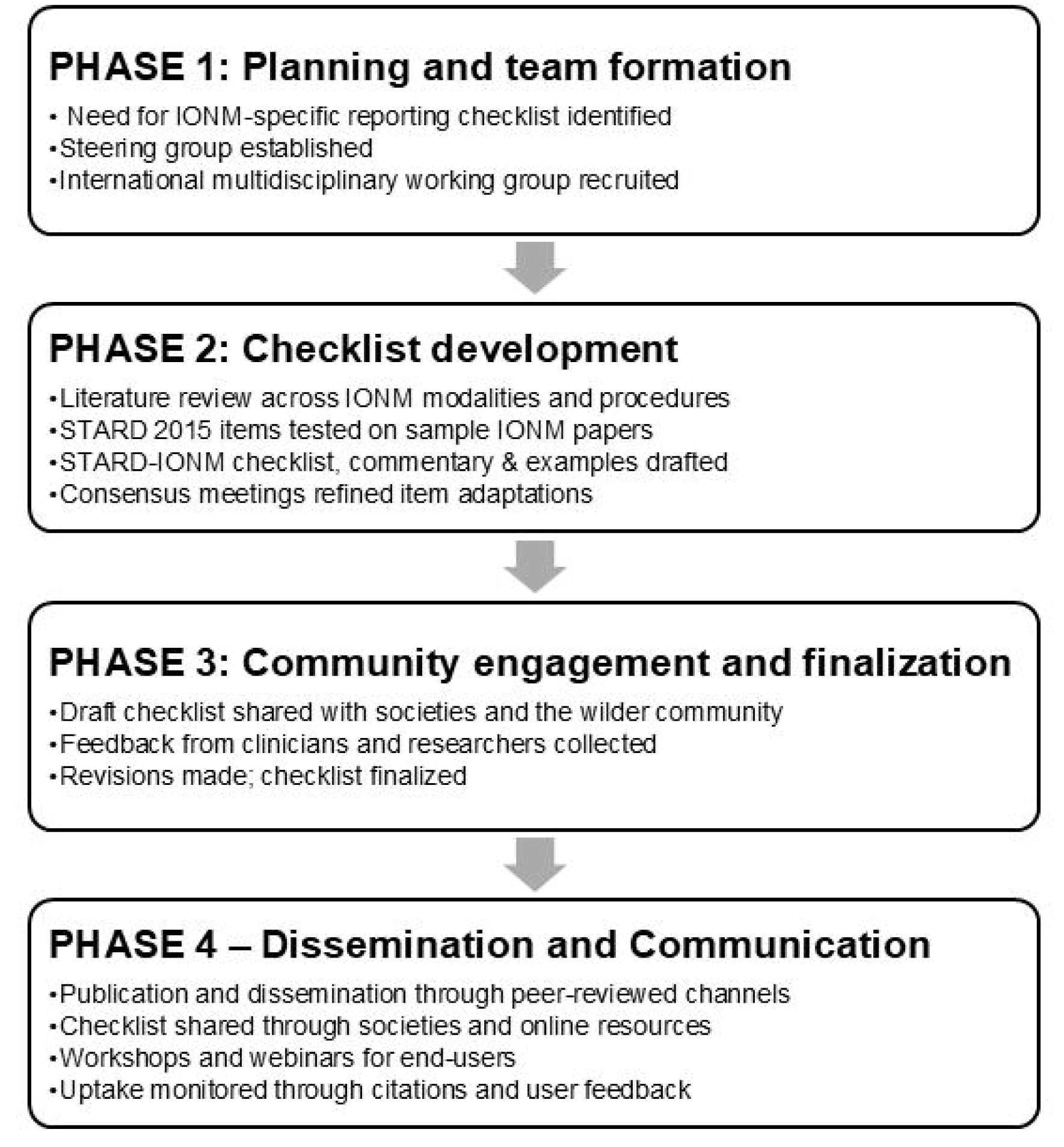
Four-phase workflow used to develop the STAED-IONM checklist. The process included planning and team formation (Phase 1), checklist development (Phase 2), community engagement and finalization (Phase 3), and dissemination (Phase 4).

### Phase 1 – Planning and Team Formation

During Phase 1, we developed the rationale for adapting the STARD framework to IONM through discussions with experts on IONM modalities. Under the guidance of PM Bossuyt, a steering group was established to oversee the initiative and coordinate efforts with the broader working group. Steering-group members were selected for their complementary expertise in clinical neurophysiology (PDT, AMH, AS, GD, JRB), neurosurgery (KS), and diagnostic-accuracy methodology (PMB), with attention to geographic diversity and gender balance. Working-group members were nominated through the steering group’s professional networks based on recognized expertise in IONM modalities, surgical domains, or clinical neurophysiology research, ensuring representation from Central, South, and North America, Europe, and Asia. Two invited members declined participation. The consensus process used a structured, iterative review rather than a formal Delphi approach. For each development task (scope definition, item evaluation, checklist drafting, and commentary writing), members first worked independently, followed by virtual discussion, written review between meetings, and version-controlled document revision. Consensus was reached through iterative independent review, virtual discussion, written feedback, and version-controlled revision. The process did not use formal voting, Delphi rounds, or prespecified decision thresholds; consensus was operationally defined as the absence of sustained objection after iterative review.

### Phase 2 – Development of STARD-IONM Checklist

Phase 2 focused on adapting the STARD-2015 checklist to the specific context of IONM. The development process included:

#### a. Scope Definition

The steering and working groups collaboratively defined the scope of the STARD-IONM extension through an extensive literature review of diagnostic accuracy studies involving IONM. We pre-specified target populations, procedures, modalities, and reference-standard domains to ensure generalizability.

#### b. Evaluation of Item Applicability

A scoping review of the IONM diagnostic-accuracy literature was first performed to characterize prevailing reporting practices and identify common gaps.

Each steering-group member nominated three candidate studies, yielding 18 nominations. Using predefined inclusion and exclusion criteria, the steering group selected 12 studies by consensus to ensure representation across IONM modalities, surgical domains, age groups, and geographic regions. The final sample included spine, cranial/skull base, vascular, and other neurosurgical applications; adult and pediatric populations; and studies from North America, Europe, and Asia. The 12 evaluated studies are listed in Supplementary Table 3.

For the STARD assessment, studies were assigned to working-group members such that each of the 12 studies was independently evaluated by three reviewers. Reviewers assessed all 30 STARD-2015 items using the following categories: “Yes,” fully and unambiguously reported; “No,” not reported; and “Unclear,” partially or ambiguously reported. Percent agreement among the three reviewers was calculated for each item. Formal inter-rater reliability statistics (e.g. kappa coefficients) were not computed for this scoping evaluation.

#### c. Development of a preliminary STARD-IONM Checklist, Commentary and Examples

Each member was assigned four STARD-2015 items for development of the STARD-IONM checklist, commentary, and examples. For each item, members prepared a brief narrative, describing its specific relevance to IONM and identified an illustrative example from a published IONM study.

#### d. Methodological Considerations Informing Checklist Adaptation

During checklist development, the steering group and working group jointly identified recurring methodological challenges that influence diagnostic-accuracy inference in IONM yet do not align with a single STARD-2015 item. In addition to providing IONM-specific commentary, the development process adopted selected STARD-2015 parent items to incorporate explicit IONM-specific reporting elements where the original checklist did not adequately capture key methodological features of IONM diagnostic-accuracy studies. Because these challenges span multiple reporting domains and can introduce bias, outcome misclassification, and limited interpretability when insufficiently specified, the working group was subsequently organized into focused subgroups aligned to member expertise (e.g., reference standards, reversibility/partial recovery, anesthesia/physiology reporting, and evaluation frameworks). These subgroups then discussed, synthesized, and drafted concise guidance paragraphs and reporting recommendations to clarify terminology, improve consistency, and provide a practical reference for authors. The resulting guidance text was refined through group deliberation and incorporated into the manuscript as a companion to the STARD-IONM checklist to support its application across diverse IONM study designs.

### Phase 3 – Community Engagement and Finalization

Phase 3 focused on engaging the broader IONM community and finalizing the checklist. A draft of the STARD-IONM reporting framework was shared with professional societies and clinical experts in neurology, neurosurgery, orthopedic surgery, anesthesiology, and clinical neurophysiology. It was also shared via professional social media platforms to invite input from clinicians and researchers involved in IONM. Proposed revisions were synthesized and resolved by the steering group through consensus and version-controlled drafts, culminating in a finalized checklist, definitions, and curated examples.

### Phase 4 - Dissemination

Phase 4 will focus on disseminating the final STARD-IONM checklist to facilitate broad adoption and to support its use as a reference standard for diagnostic accuracy reporting in IONM through multiple channels.

## Results

### Phase 1. Steering-group formation to define rationale and planning

We established a six-member steering group—PDT, AMH, JRB, AS, KS, GD—to define scope, governance, and early deliverables. This team brought together expertise in clinical neurophysiology, neurosurgery, and diagnostic-accuracy methodology and achieved cross-regional and gender-diverse representation. In parallel, an 18-member working group was constituted with contributors from Central, South and North America, Europe, and Asia, ensuring worldwide representation for item review and consensus input. Phase-1 outputs were: (i) consensus to develop an IONM-specific extension of STARD-2015; (ii) agreement on a four-phase program; (iii) role assignments aligned to CRediT (Contributor Roles Taxonomy); and (iv) confirmation of funding and disclosures—thereby laying the foundation for checklist drafting and community engagement in subsequent phases.

### Phase 2. Scope definition, evaluation of item applicability and draft STARD-IONM checklist

The outcomes of Phase 2 include (a) scope definition; (b) structured analysis of item-level applicability; (c) development of a preliminary STARD-IONM checklist and identification of high-quality examples of IONM reporting; and (d) developing cross-cutting guidance on recurring methodological challenges via expert subgroups and consensus refinement. These results informed the refinement of the STARD-IONM checklist and the accompanying guidance document.

#### a. Scope Definition Achieved

Consensus domains for reporting diagnostic accuracy studies in IONM emerged from steering- and working-group deliberations. The final sample comprised 12 studies spanning adult and pediatric populations, single- and multicenter designs, and diverse geographies (North America, Europe, Asia, and additional regions).

– Clinical coverage: Surgical contexts were broadly represented: spine/spinal cord; intracranial procedures (eloquent and non-eloquent cortex); neurovascular/cerebrovascular surgery; peripheral nerve; and skull base operations.
– IONM modalities: Index tests reflected routine practice: SSEP and transcranial/direct-cortical MEP were most frequent, with additional use of EMG (free-run/triggered), EEG/ Electrocorticography, BAEP, visual evoked potentials, and cortico-cortical evoked potentials (CCEPs).
– Reference standards: Studies predominantly used postoperative clinical outcome scales, with surrogate end points where appropriate (e.g., cerebral ischemia, pedicle-screw placement accuracy).
– Contextual factors: Studies reported a range of anesthesia techniques and physiological conditions during monitoring, and reference standards were applied at varying time points (immediate postoperative, discharge, and longer follow-up). Definitions of the index test and target condition also differed across surgical contexts.
– Summary: Collectively, these findings delineate typical IONM use in diagnostic-accuracy research across procedures, modalities, and outcomes, and they informed the domain structure and examples included in Table 1.

**Table 1.** Scope Domains Defined in Phase 2 of the STARD-IONM Initiative.

| Scope Domain | Definition |
| --- | --- |
| Populations and settings | Adult; pediatric; single- vs multi-center; North America / Europe / Asia / other regions. |
| Target study designs | Diagnostic accuracy studies evaluate IONM performance in detecting or predicting intraoperative neurological events or postoperative outcomes. |
| Surgical applications | Procedures where IONM assesses neural integrity or guides decisions: spine/spinal cord, intracranial (eloquent/non-eloquent), neurovascular/cerebrovascular, peripheral nerve, skull base. |
| IONM modalities | Somatosensory Evoked Potentials (SSEPs), transcranial and direct cortical MEPs, EEG, IOM-EMG, Brainstem Auditory Evoked Potentials (BAEPs), Visual Evoked Potentials (VEPs), and cortico-cortical evoked potentials. |
| Anesthesia and physiological context | General anesthesia and total intravenous anesthesia (TIVA), considering their effect on IONM signal interpretation. |
| Reference standards | Clinical outcomes (e.g., postoperative motor/language/cranial nerve deficits), surrogate outcomes (e.g., cerebral ischemia, effectiveness of microvascular decompression using lateral spread response and pedicle screw placement accuracy). |
| Reference-standard timing | Timing of outcome assessment, including immediate postoperative evaluation, assessment at discharge, and longer-term follow-up, depending on the target condition and IONM modality. |

**Table 2:**
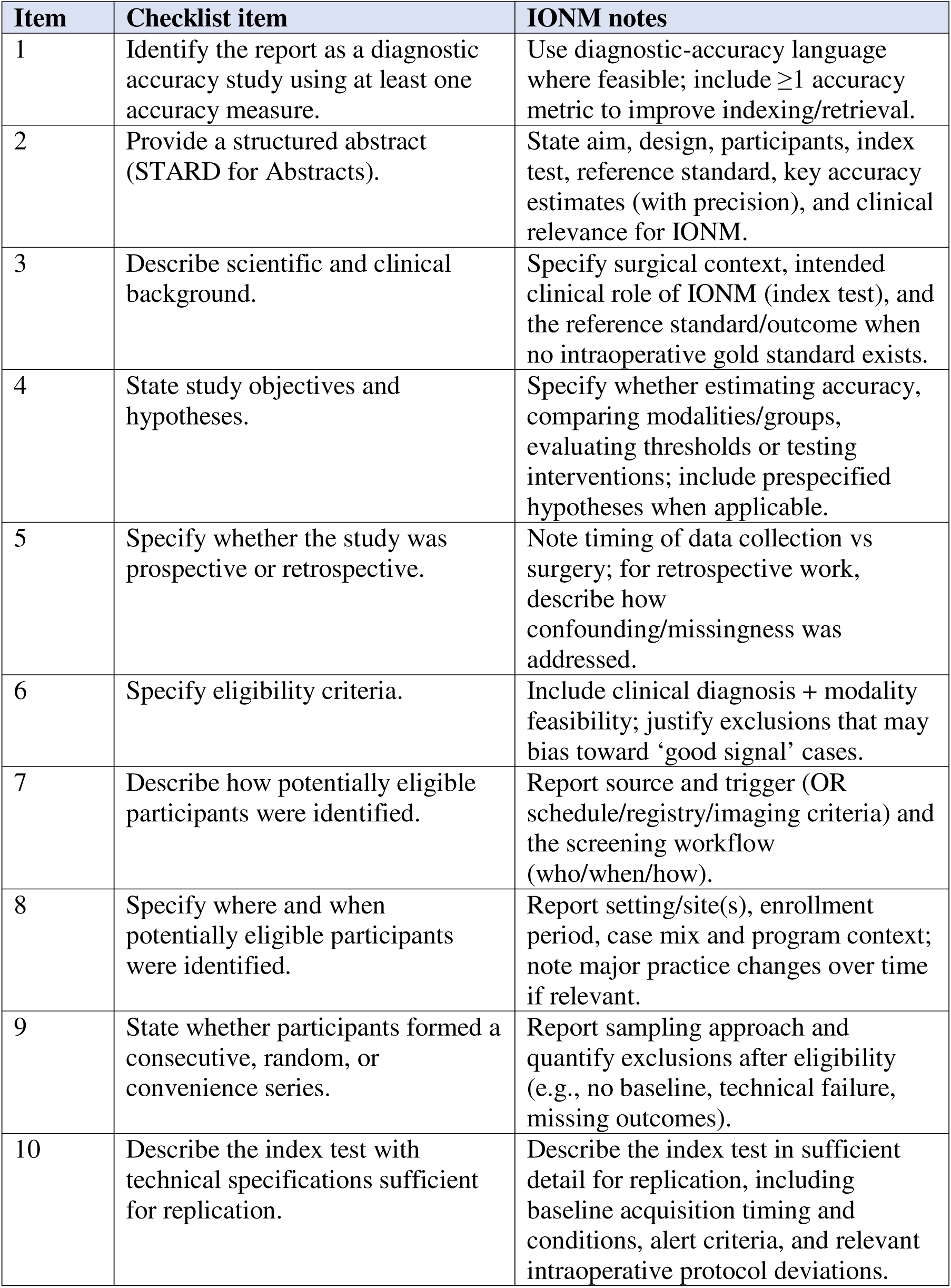
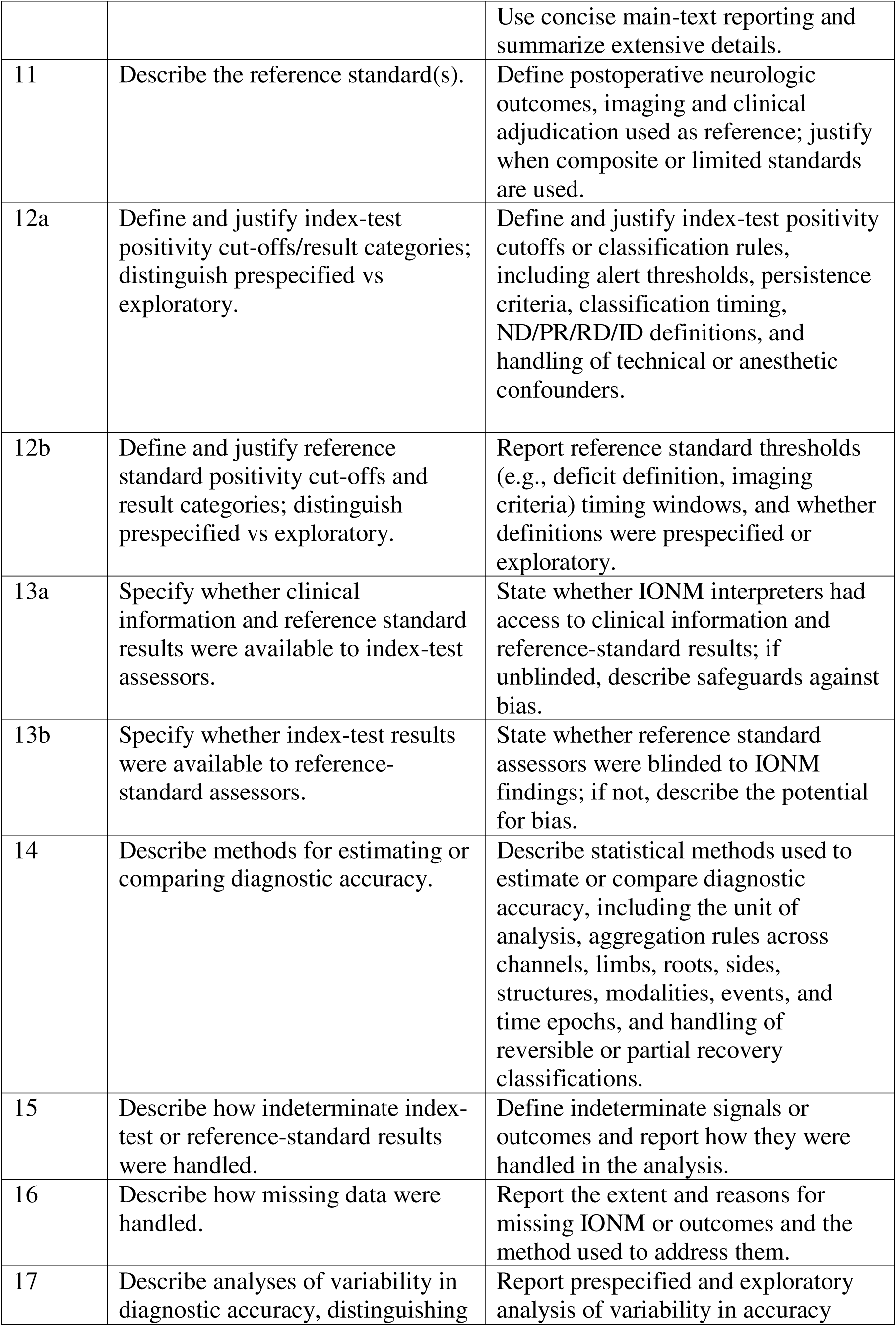
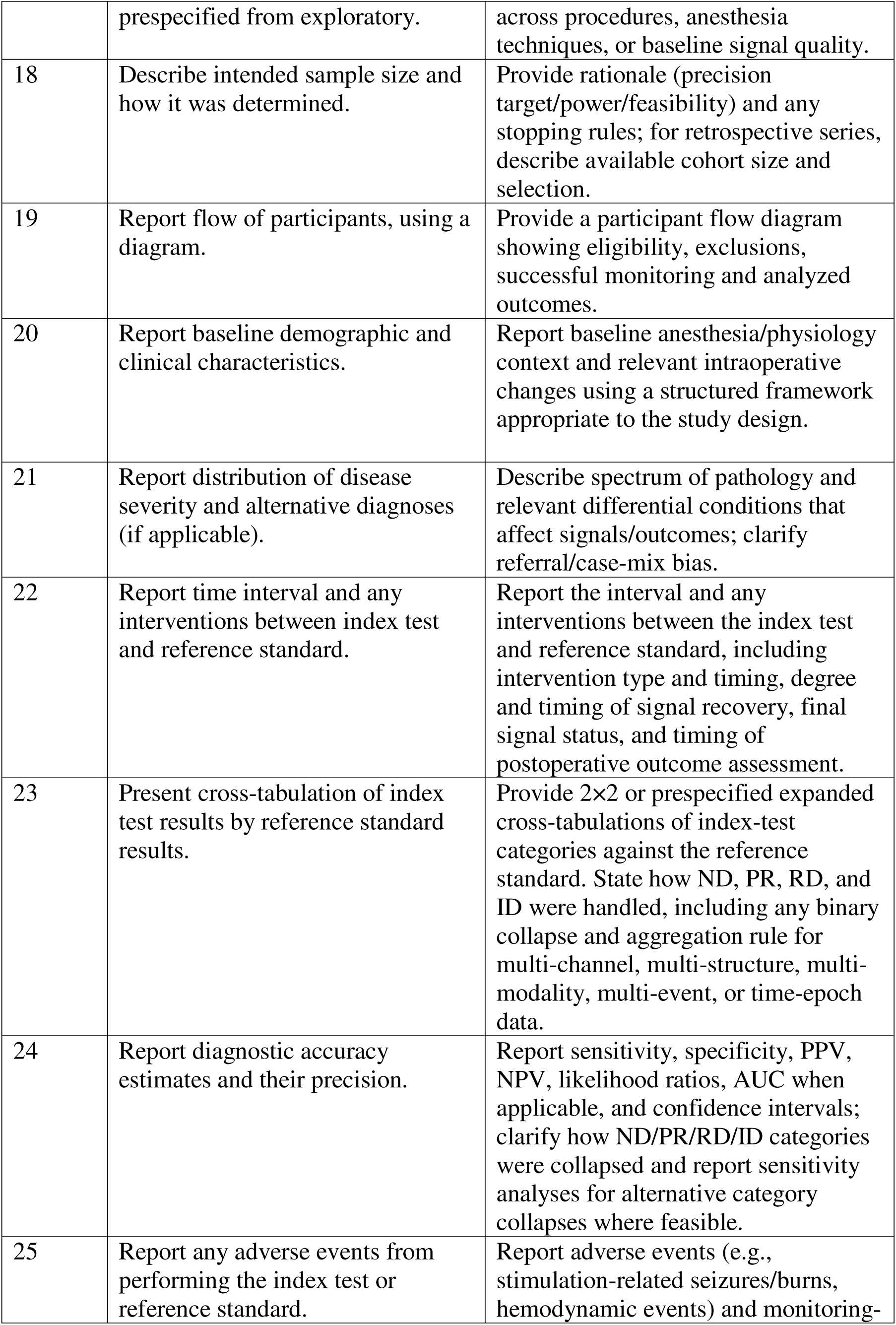
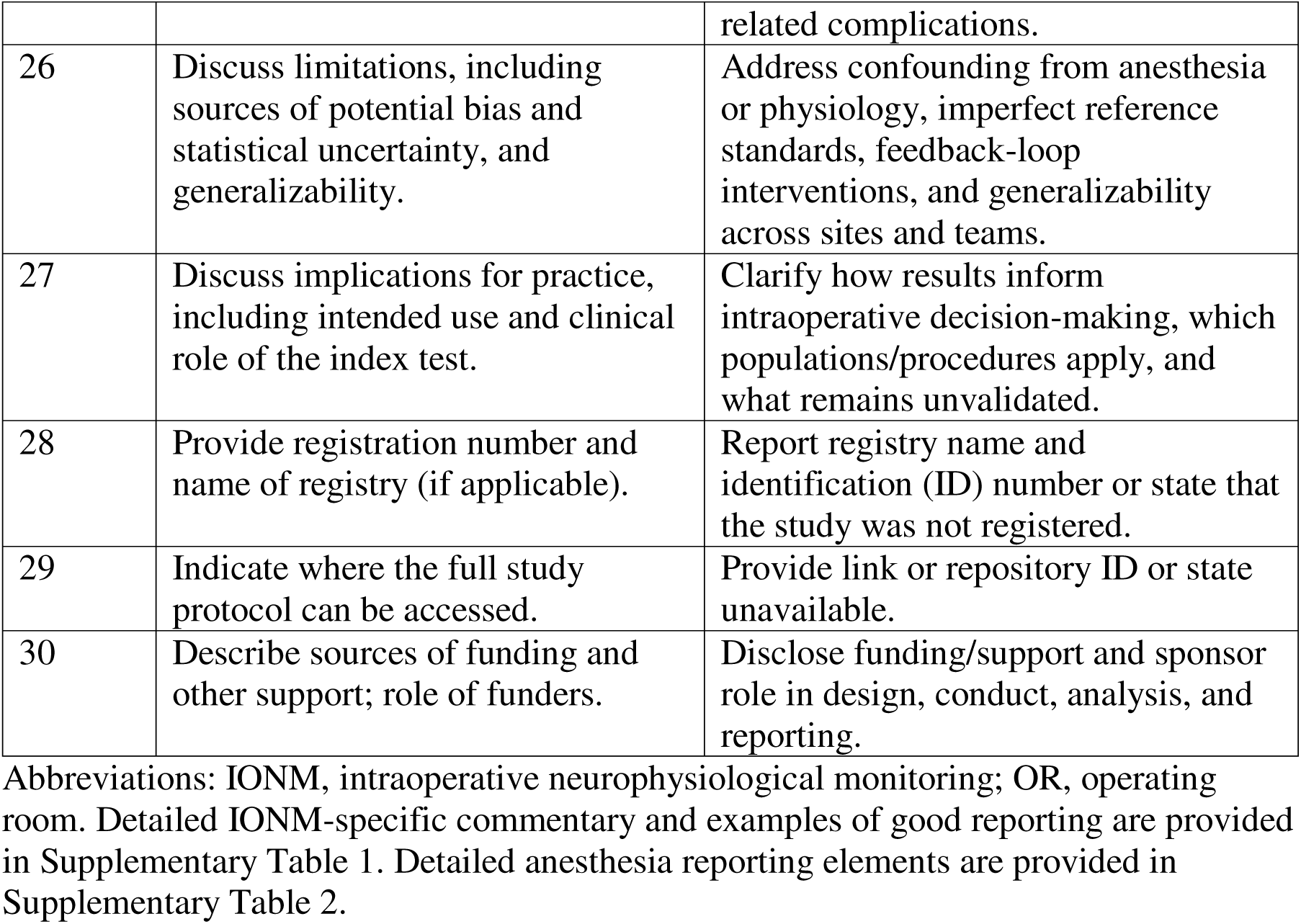
STARD-IONM checklist. Columns: Item | STARD-IONM Checklist item | IONM-specific notes

#### b. Evaluation of item Applicability

Results (Figure 2) showed that item-level reporting revealed substantial variability. Percent agreement was used descriptively to assess scoring consistency in this scoping evaluation. Agreement varied across checklist items, with disagreement occurring most often when source studies reported items incompletely or ambiguously rather than when scoring definitions were unclear. These observations informed prioritization of IONM-specific commentary in the checklist. Frequently, foundational elements were included, such as the scientific background and rationale for the index test (Item 3: 27/30, 90%) and discussion of clinical applicability (Item 27: 26/30, 86%)). In contrast, key methodological elements were underreported, such as the handling of missing data (Item 16: 2/30, 6.7%), adverse events from index test or reference standard (Item 25: 3/30, 10%), and availability of study protocols (Item 29: 1/30, 3.3%). Reviewer agreement was consistent for 21/30 (70%) items, while 9/30 (30%) items showed mixed ratings, indicating ambiguity or insufficient methodological detail.

**Figure 2:**
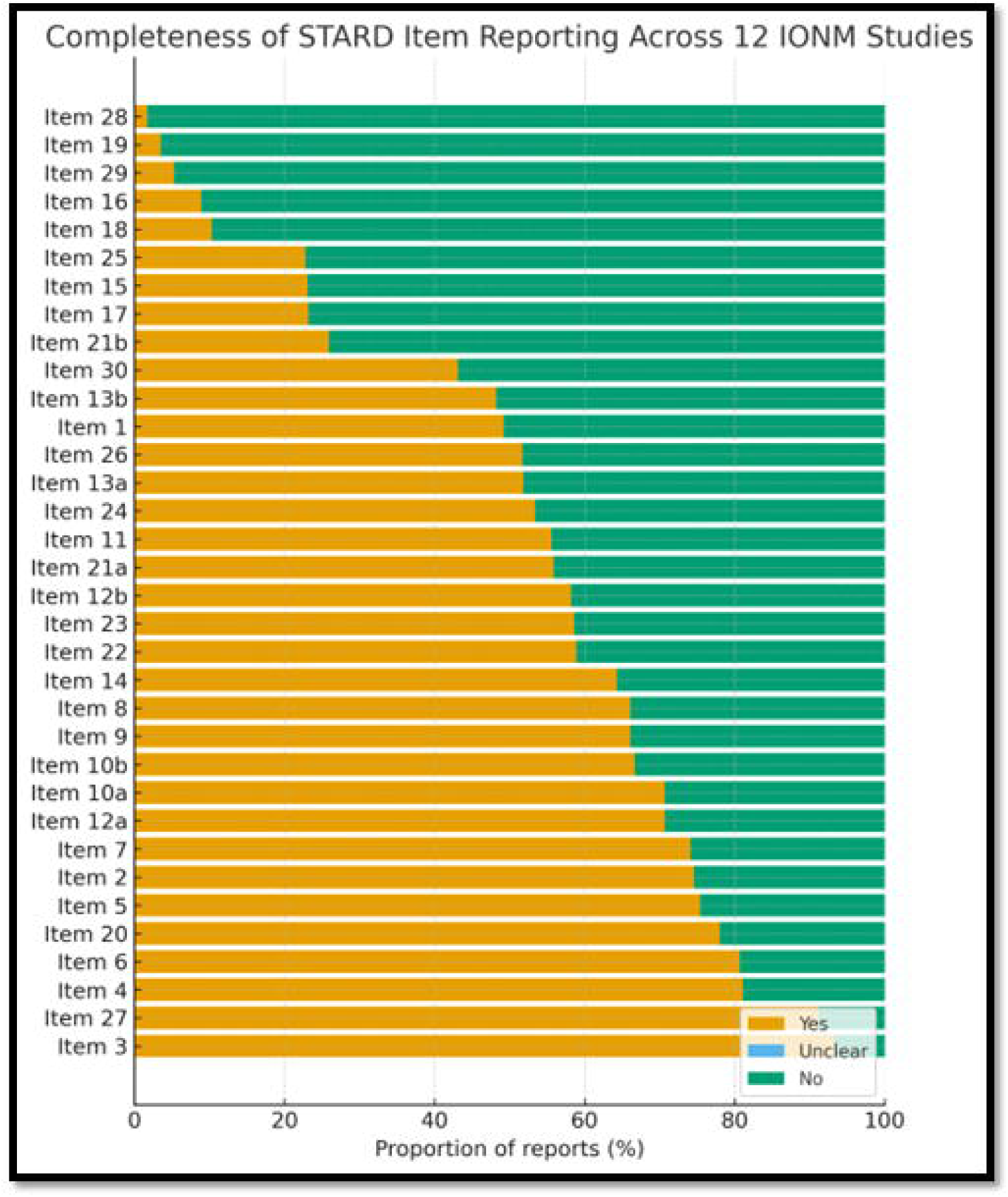
Completeness of STARD-2015 item reporting across 12 IONM studies. Stacked bars show, for each of the 30 STARD-2015 checklist items, the distribution of summarized item-level ratings across the 12 evaluated studies, classified as Yes, Unclear, or No. Items are ordered by the proportion rated Yes. The figure highlights variability in reporting completeness across checklist items and identifies underreported methodological domains

#### c. Development of preliminary STARD-IONM Checklist, Commentary and Examples

The final STARD-IONM checklist retained the STARD-2015 framework while revising selected parent items to incorporate explicit IONM-specific reportable elements, including alert thresholds and persistence rules; signal-event classification <u>as no deterioration (ND), partial recovery (PR), reversible deterioration (RD), and irreversible deterioration (ID);</u> intervention timing and final signal status; unit of analysis and aggregation; and structured anesthesia/physiology reporting. Building on STARD 2015, the steering and working groups produced three linked outputs for each item: (1) a STARD-IONM checklist directive specifying the reportable item, (2) a brief IONM-specific commentary outlining the rationale, common pitfalls and minimum reporting elements, and (3) a published examples of adequate reporting, with PMID/DOI and brief justification. Wording was standardized and reviewed by the steering group to enhance clarity and transparency. The checklist was aligned with core principles of diagnostic accuracy reporting, with options for alternative reporting frameworks. STARD-IONM checklist specific commentary and examples of good reporting with citation are provided (Supplementary Table 1).

#### d. Methodological Consideration Informing Checklist Adaptation

The working-group sub teams produced consensus guidance across four domains: (1) reference standards (selection, implementation, timing, and blinding); (2) reversibility/partial recovery (definitions and scoring of ND/PR/RD/ID with intervention documentation); (3) anesthesia/physiology (a structured minimum reporting set); and (4) evaluation framework (threshold specification, handling of RD/PR, and metric rationale). These consensus statements were edited and incorporated as manuscript-ready guidance text to accompany the STARD-IONM checklist and support consistent application across IONM study designs.

##### 1. Reference Standards

Reference standards in IONM studies may include elements of preoperative and postoperative neurological assessments, validated clinical scales, diagnostic tests, and patient-reported outcomes. Authors must justify the rationale for selecting reference standards, and describe their implementation, timing, procedural details, and personnel qualifications to ensure consistency and reduce variability. Reference standards in IONM studies should be tailored to the operative domain and reported with sufficient detail including the assessment method, assessor qualifications, timing, positivity threshold or severity category, duration/permanence, and clinical relevance to patient-centered outcomes. Examples include standardized postoperative neurologic examination for spine/spinal cord monitoring, postoperative CT and neurologic examination for pedicle screw studies, structured cranial nerve examination for skull base procedures, formal postoperative language assessment after awake mapping, postoperative stroke or focal neurologic assessment after vascular procedures, and targeted postoperative motor/sensory examination for peripheral nerve applications. Illustrative domain-specific examples are provided in Supplementary Table 4. Reference standards should be described in sufficient detail to enable study replication. The use or absence of blinding of outcome assessment must be explicitly stated. Authors should prespecify and justify the index-test time point or aggregation rule used for comparison with the reference standard. For predictive studies, comparison with the final IONM signals recorded shortly prior to the end of surgery is appropriate. For detection-oriented studies, comparison may include transient IONM changes (alerts) regardless of subsequent recovery, and intraoperative reference standards may be used. The rationale for the chosen time point should be stated explicitly. Intraoperative reference standards (e.g. Stagnara wake up test, intraoperative angiography or neuroimaging) may be compared to transient IONM changes (a positive index test result) that later revert to normal following surgical intervention. However, certain intraoperative reference standards—such as brief neurological assessments conducted in sedated patients —are inherently prone to bias due to time constraints and limited access to the patient.

##### 2. Reversibility/ partial recovery

Reversibility of IONM changes remains a central challenge in diagnostic test accuracy (DTA) research because results are commonly categorized as no deterioration (ND), irreversible deterioration (ID), and reversible deterioration (RD). Conventional 2×2 DTA analyses typically treat ND as negative and ID as positive, but RD is not readily reducible to a binary outcome because it reflects a transient deterioration that often resolves in response to intraoperative actions (e.g., surgical adjustment) and may represent successful injury prevention via intraoperative therapeutic intervention. Reversible deterioration (RD) has been handled inconsistently across IONM diagnostic-accuracy studies, and each common approach introduces tradeoffs. Strategies include context-dependent scoring (risking incorporation bias), end-of-surgery scoring or exclusion of RD (both potentially increasing spectrum bias and reducing interpretability), and binary classification of RD as “positive,” which can misclassify successfully mitigated events as false positives. Multicategory tables preserve the full distribution and enable richer risk stratification, but do not map cleanly onto conventional 2×2 accuracy metrics. Notably, many studies do not explicitly report how RD was classified, limiting comparability and reproducibility across literature.(Clark et al., 2013; Hilibrand et al., 2004; Holdefer et al., 2016; Kobayashi et al., 2011; Wilent et al., 2020) STARD-IONM defines no deterioration (ND), partial recovery (PR), reversible deterioration (RD), and irreversible deterioration (ID) as intraoperative index-test signal-event classifications rather than postoperative outcome categories. These classifications describe the intraoperative signal course relative to prespecified alert, persistence, recovery, and classification-time criteria, whereas postoperative neurological status remains the reference standard. ND indicates that no prespecified alert threshold was crossed. RD indicates that a prespecified alert threshold was crossed, technical and anesthetic explanations were reasonably excluded, and the signal returned to within prespecified recovery criteria by the prespecified classification time point. PR indicates that the signal improved after crossing the alert threshold but did not meet full-recovery criteria by the prespecified classification time point. ID indicates that signal loss or deterioration persisted at the prespecified classification time point. For each alert, authors should report baseline timing and conditions, the modality or structure involved, the channel, limb, root, side, vascular territory, or monitored structure affected, the alert threshold crossed, the persistence rule used to define an alert, the classification time point, technical, anesthetic, and physiologic causes considered and excluded, the intervention type and timing, the degree and timing of recovery, the final signal status, and the timing and method of postoperative outcome assessment. Authors should prespecify the unit of analysis and aggregation rules before estimating diagnostic accuracy. These rules should state how multiple channels, limbs, roots, sides, structures, modalities, alerts, and time epochs are combined into patient-level, structure-level, event-level, or epoch-level classifications.

##### 3. Anesthesia/ Physiology

Standardized reporting of anesthesia-related variables is essential for interpreting IONM signals and ensuring study reproducibility. The STARD-IONM initiative (Supplementary Table 2) suggests documentation in the core domains: patient demographics; anesthetic technique; depth of anesthesia monitoring; neuromuscular blockade; physiological monitoring; intraoperative physiological management; and communications and actions. To improve feasibility and adoption, the STARD-IONM anesthesia and physiology reporting framework (Supplementary Table 2) incorporates minimum, enhanced, and ideal prospective reporting elements within each domain. This framework provides a structured approach to consistent documentation of variables known to influence IONM signal quality and patient outcomes and serves as a companion to the broader STARD-IONM checklist.

##### 4. Evaluation framework

The evaluation of diagnostic accuracy in IONM poses unique challenges due to the dynamic and partially reversible nature of IONM signal changes. Although conventional metrics such as sensitivity, specificity, and 95% confidence intervals are commonly reported,(Holdefer et al., 2013) their application in IONM studies requires careful methodological consideration. Receiver operating characteristic (ROC) curve analysis is frequently employed to determine optimal thresholds for EP alerts.(Muramoto et al., 2013) However, thresholds derived post hoc from ROC analysis (as compared to planned thresholds) may introduce bias.(Pan et al., 2024) Authors should clearly state whether thresholds were prespecified or derived from the data and discuss the implications for diagnostic accuracy. As outlined in Section b, the classification of reversible deterioration (RD) significantly impacts diagnostic accuracy calculations. Yet many studies fail to report how recovered EP changes were classified, limiting transparency and reproducibility. To address this, the STARD-IONM checklist recommends explicit documentation of : a) the scoring method used for EP changes, including RD and PR; b) whether alert thresholds were prespecified or derived from ROC analysis; c) the rationale for chosen evaluation metrics and their methodological limitations.

Adopting standardized and transparent evaluation frameworks will allow IONM research to improve methodological rigor, facilitate cross-study comparisons, and enhance its contribution to evidence-based surgical neurophysiology.

### Phase 3. Community Engagement

In Phase 3, feedback was sought from a broad range of IONM stakeholders, including members of professional societies in neurology, neurosurgery, orthopedic surgery, anesthesiology, and clinical neurophysiology, as well as clinicians and researchers reached through professional social media. Responses were received from individuals across these groups, and no concerns were raised regarding the overall scope of the checklist. Comments primarily focused on improving clarity, operational definitions, and reporting feasibility, particularly with respect to signal-recovery categories, the handling of reversible changes in diagnostic-accuracy analyses, anesthesia and physiology reporting, the timing of key intraoperative recordings, and blinding and outcome assessment. Several of these issues had already been identified during earlier rounds of iterative review within the geographically diverse working group, prior to external circulation. The main revisions resulting from this input were the addition of partial recovery as an explicit signal-change category, refinement of the anesthesia and physiology framework to incorporate minimum, enhanced, and ideal prospective elements within each domain, expansion of guidance on blinding, and language edits to improve consistency and clarity. The final STARD-IONM checklist and accompanying guidance were then completed through version-controlled revision and steering-group review.

### Phase 4 – Dissemination of STARD-IONM checklist

The finalized checklist and guidance will be disseminated via (i) peer-reviewed publication with open-access hosting of the checklist and supplements; (ii) targeted outreach to professional societies in neurology, neurosurgery, orthopedics, anesthesiology, and clinical neurophysiology; and (iii) community engagement through workshops, scientific sessions, and academic social media to support visibility and adoption in IONM research and peer review. Following publication, all authors will be encouraged to present and discuss STARD-IONM at regional, national, and international meetings focused on IONM and perioperative neuroscience. This author-led dissemination will complement society partnerships and online campaigns, reinforcing consistent messaging and practical uptake in study design, reporting, and editorial practice. Future dissemination efforts may also explore the development of AI-assisted manuscript evaluation tools that leverage the STARD-IONM checklist, item-level commentary, and curated examples (Supplementary Table 1) to support authors in identifying underreported items and improving reporting completeness during manuscript preparation.

## Discussion

This paper introduces the STARD-IONM checklist, a tailored extension of the STARD 2015 framework, developed to improve the reporting of diagnostic accuracy in IONM. Created through a structured consensus process and broad community engagement, the checklist addresses key methodological gaps. As IONM becomes increasingly integrated into complex surgical workflows, its role in guiding intraoperative decision-making is more critical than ever. This underscores the need to establish a consensus on consistent and reliable reporting standards. However, both existing and future IONM studies face challenges that affect their reliability and interpretability. These include the subjective interpretation of neurophysiological changes, non-standardized definitions of “significant” signal alterations, and variability in reference standards, all of which complicate the assessment of IONM’s diagnostic performance. Additionally, intraoperative anesthetic and surgical interventions may modify patient outcomes, making it difficult to distinguish true from false positives, an essential step in determining IONM test accuracy. Inconsistent definitions, timing, and reporting of outcome assessments further obscure the evidence base and limit its clinical utility.

A key challenge in evaluating IONM is its dual role not only as a diagnostic test, but also as a trigger for real-time therapeutic interventions. IONM diagnostic accuracy studies may address fundamentally different questions, each requiring a distinct analytic framework: (1) Prediction: Does the final IONM signal status predict postoperative neurological outcome? This question compares end-of-surgery signal classification (ND/PR/RD/ID) with the reference standard and is most directly aligned with conventional diagnostic accuracy analysis. (2) Detection: Does IONM reliably detect an intraoperative neurological threat at the time it occurs? This question evaluates whether IONM alerts correspond to surgically or physiologically meaningful events, regardless of whether the outcome is ultimately favorable due to intervention. The appropriate reference standard may be an intraoperative comparator (e.g., angiography, wake-up test) or a composite of intraoperative events and postoperative outcomes. (3) Intervention effectiveness: Does acting on IONM alerts improve outcomes compared with not acting? This question requires a different study design altogether—ideally a controlled comparison of outcomes with and without IONM-guided intervention—and is not well served by standard sensitivity/specificity analysis. Authors should explicitly state which question their study addresses, as the choice determines the appropriate index-test time point, reference standard, and analytic approach. The interventional nature of IONM creates important challenges for the interpretation of diagnostic accuracy.

Interventions prompted by IONM alerts can alter the course of surgery and affect outcomes, yet are rarely described in sufficient detail. This lack of intervention reporting complicates the interpretation of test accuracy and highlights a fundamental limitation of applying standard STARD criteria to IONM. In the literature, significant changes that recover after intervention have been classified inconsistently as either positive or negative test results.(Holdefer and Skinner, 2020) Moreover, when interventions reverse abnormalities without a clear reference standard, it becomes difficult to determine whether the outcome represents a true or false positive. These issues underscore the complexity of IONM and the need for tailored reporting standards that reflect its dynamic and interventional nature. For example, during a carotid endarterectomy, a sudden EEG change prompts the team to insert a shunt, the EEG promptly recovers, and the patient wakes without neurological deficit. Without an independent reference standard, it remains unclear whether the alert was a true positive (averted ischemia) or a false positive related to anesthetic or physiological factors.

STARD-IONM is designed as a practical reporting framework applicable across diverse IONM research contexts. In primary studies, investigators introducing new IONM modalities should clearly define the index test, prespecified alert thresholds, and reference standards for postoperative outcomes. For investigators reporting studies on existing IONM modalities, STARD-IONM emphasizes diagnostic accuracy and reliability reporting, including precision estimates, and approach to interpretation of reversible and irreversible signal changes. In systematic reviews, STARD-IONM supports consistent data abstraction by clarifying terminology and outcome definitions. While sensitivity and specificity are not fixed test properties, as explicitly noted in STARD 2015, complete reporting that enables their calculation remains a necessary first step. Once basic transparency is achieved, more nuanced metrics can be developed to better reflect the complexity of IONM.(Bossuyt et al., 2015)

STARD-IONM provides a structured framework to translate improved reporting into more reliable interpretation and application of IONM research. By promoting clarity, completeness, and consistency in the documentation of index tests, reference standards, alert criteria, recovery patterns, and intraoperative interventions, the checklist strengthens the foundation of diagnostic accuracy studies in IONM. Improved reporting supports more rigorous appraisal and evidence synthesis, facilitates meaningful synthesis of evidence across studies, and enhances comparability across modalities, procedures, and institutions. While not all studies may fulfill every STARD-IONM checklist item, authors are encouraged to transparently justify any omissions. Beyond guiding researchers, STARD-IONM also offers a practical tool for peer reviewers and journal editors to evaluate the quality and completeness of IONM research, fostering greater reproducibility and interpretability across the field.

A limitation of the development process is that patient, family, and public involvement was not incorporated into the development of the STARD-IONM checklist. Future revisions should consider including these perspectives, particularly regarding patient-reported outcomes, patient-centered reference standards, and the clarity and relevance of reported outcomes.

## Conclusion

The STARD-IONM extension builds on the STARD-2015 framework to provide item-level reporting standards tailored to the unique context of IONM. By clarifying reference standards, documenting reversible changes along with related alerts and interventions, and addressing anesthesia-related confounders, the checklist directly responds to the recurrent gaps identified. We recommend adoption and endorsement of STARD-IONM by authors, reviewers, editors, professional societies, and journals to support consistent and transparent reporting. Its implementation is expected to improve reporting completeness, facilitate evidence synthesis, and strengthen the foundation for evidence-based IONM research.

## Supporting information

Supplementary Table 3 Publications for ITEM evaluation

## Data availability

The STARD-IONM checklist, supplementary tables, audit materials, and the list of evaluated studies will be deposited in a permanent public repository upon acceptance. A DOI-linked repository URL will be provided in the final published version. Until then, all materials are available as supplementary files accompanying this manuscript.

## CRediT authorship contribution statement

Conceptualization: PDT, GD, JRB, AS, AMH, KS, PB.

Methodology: PDT, PB, GD.

Investigation/Data curation: All authors.

Formal analysis: All authors.

Writing – original draft: PDT, GD.

Writing – review & editing: All authors.

Visualization: PDT.

Supervision/Project administration: PDT.

Funding acquisition: AMH

Guarantor(s): PDT, GD.

## Acknowledgements

We thank the members of the **STARD-IONM Working Group** for item-level evaluation, discussion, and consensus input during development of the checklist.

## STARD-IONM Working Group

Anthony R. Absalom^7^, Stefanie Binzer^8^, Michael G. Fehlings^9^, Isabel Fernandez-Conejero^10^, Lanjun Guo^11^, Robert Holdefer^12^, Ernest M. Hoffman^13^, David B. McDonald^14^, Marc R. Nuwer^15^, Kyung Seok Park^16^, Julian Prell^17^, Francesco Sala^18^, Nishanth Sampath^19^, Daniel San-Juan^20^, Jay L. Shils^21^, Mirela V. Simon^22^, Christoph N. Seubert^23^, Silvia M. Verst^24^

^7^ Department of Anesthesiology, University Medical Center Groningen, Groningen, Netherlands

^8^ Department of Neurology, Odense University Hospital, Denmark

^9^ Division of Neurosurgery, University of Toronto, Toronto, ON, Canada

^10^ Head of Intraoperative Neurophysiology Unit, University Hospital of Bellvitge (Barcelona), Spain

^11^ Department of Neurosurgery, University of California San Francisco (UCSF), CA, USA

^12^ Department of Neurology, University of Washington, Seattle, WA, USA

^13^ Department of Neurology, Mayo Clinic, Rochester, MN, USA

^14^ Advisor and past President, International Society of Intraoperative Neurophysiology; Scientific Director, Arkana Forum

^15^ Department of Neurology, UCLA, Los Angeles, CA, USA

^16^ Department of Neurology, Seoul National University Bundang Hospital, Seoul National University College of Medicine, Seoul, Korea

^17^ Department of Neurosurgery, Universitätsklinikum Halle (Saale), Martin Luther University Halle-Wittenberg, Halle (Saale), Germany

^18^ Department of Neurosurgery, University of Verona, Verona, Italy

^19^ Department of Clinical Neurophysiology, Institute of Neurosciences, SIMS Hospital, Chennai, India

^20^ Epilepsy Clinic, National Institute of Neurology and Neurosurgery, Mexico City, Mexico

^21^ Department of Neurosurgery, Rush University Medical Center, Chicago, IL, USA

^22^ Department of Neurology, Beth Israel Deaconess Medical Center, Boston, MA, USA

^23^ Envision Physician Services, HCA Ocala; Department of Anesthesiology, University of Central Florida, Orlando, FL, USA

^24^ IOM Program Coordinator of the Clinical Neurophysiology Residency at University of São Paulo Medical School, São Paulo, Brazil

## References

Asimakidou E, Abut PA, Raabe A, Seidel K. Motor Evoked Potential Warning Criteria in Supratentorial Surgery: A Scoping Review. Cancers (Basel) 2021;13. 10.3390/cancers13112803.

Bossuyt PM, Reitsma JB, Bruns DE, Gatsonis CA, Glasziou PP, Irwig L, et al. STARD 2015: an updated list of essential items for reporting diagnostic accuracy studies. BMJ 2015:h5527. 10.1136/bmj.h5527.

Bossuyt PM, Reitsma JB, Bruns DE, Gatsonis CA, Glasziou PP, Irwig LM, et al. Towards Complete and Accurate Reporting of Studies of Diagnostic Accuracy: The STARD Initiative. Ann Intern Med 2003;138:40. 10.7326/0003-4819-138-1-200301070-00010.

Chang R, Reddy RP, Sudadi S, Balzer J, Crammond DJ, Anetakis K, et al. Diagnostic accuracy of various EEG changes during carotid endarterectomy to detect 30-day perioperative stroke: A systematic review. Clin Neurophysiol 2020;131:1508–16. 10.1016/j.clinph.2020.03.037.

Clark AJ, Ziewacz JE, Safaee M, Lau D, Lyon R, Chou D, et al. Intraoperative neuromonitoring with MEPs and prediction of postoperative neurological deficits in patients undergoing surgery for cervical and cervicothoracic myelopathy. Neurosurg Focus 2013;35:E7. 10.3171/2013.4.FOCUS13121.

Cohen JF, Korevaar DA, Altman DG, Bruns DE, Gatsonis CA, Hooft L, et al. STARD 2015 guidelines for reporting diagnostic accuracy studies: explanation and elaboration. BMJ Open 2016;6:e012799. 10.1136/bmjopen-2016-012799.

Deletis V, Shils JL, Sala F, Seidel K, editors. Neurophysiology in Neurosurgery: A Modern Approach. 2nd ed. Elsevier; 2020. 10.1016/B978-0-12-815000-9.00045-9.

Duffau H. Stimulation mapping of white matter tracts to study brain functional connectivity. Nat Rev Neurol 2015;11:255–65. 10.1038/nrneurol.2015.51.

Dulfer SE, Gadella MC, Sahinovic MM, Lange F, Absalom AR, Groen RJM, et al. Stimulation parameters for motor evoked potentials during intraoperative spinal cord monitoring. A systematic review. Clin Neurophysiol 2023;149:70–80. 10.1016/j.clinph.2023.02.170.

Dulfer SE, Groen H, Groen RJM, Absalom AR, Sahinovic MM, Drost G. The Association of Physiological and Pharmacological Anesthetic Parameters With Motor-Evoked Potentials: A Multivariable Longitudinal Mixed Model Analysis. Anesth Analg 2024;139:609–16. 10.1213/ANE.0000000000006757.

Dulfer SE, Groen RJM, Lange F, Tamasi K, Wapstra FH, Mennink LM, et al. The Influence of Vasopressor-Induced Arterial Blood Pressure Elevation on Muscle-Recorded Motor Evoked Potentials. Anesth Analg 2025. 10.1213/ANE.0000000000007701.

Gerritsen JKW, Karschnia P, Bello L, Hervey-Jumper S, Young JS, Seidel K, et al. A comprehensive framework for glioma surgery by the PIONEER Consortium and RANO resect group, part 1: intraoperative recommendations for mapping, monitoring, and decision making. Lancet Oncol 2026;27:e11–23. 10.1016/S1470-2045(25)00531-5.

Hilibrand AS, Schwartz DM, Sethuraman V, Vaccaro AR, Albert TJ. Comparison of transcranial electric motor and somatosensory evoked potential monitoring during cervical spine surgery. J Bone Joint Surg Am 2004;86:1248–53. 10.2106/00004623-200406000-00018.

Holdefer RN, Heffez DS, Cohen BA. Utility of Evoked EMG Monitoring to Improve Bone Screw Placements in the Cervical Spine. J Spinal Disord Tech 2013;26:E163–9. 10.1097/BSD.0b013e31828871a1.

Holdefer RN, MacDonald DB, Guo L, Skinner SA. An evaluation of motor evoked potential surrogate endpoints during intracranial vascular procedures. Clin Neurophysiol 2016;127:1717–25. 10.1016/j.clinph.2015.09.133.

Holdefer RN, Skinner SA. Motor evoked potential recovery with surgeon interventions and neurologic outcomes: A meta-analysis and structural causal model for spine deformity surgeries. Clin Neurophysiol 2020;131:1556–66. 10.1016/j.clinph.2020.03.024.

Kawaguchi M, Iida H, Tanaka S, Fukuoka N, Hayashi H, Izumi S, et al. A practical guide for anesthetic management during intraoperative motor evoked potential monitoring. J Anesth 2020;34:5–28. 10.1007/s00540-019-02698-2.

Kobayashi M, Ogasawara K, Yoshida K, Sasaki M, Kuroda H, Suzuki T, et al. Intentional hypertension during dissection of carotid arteries in endarterectomy prevents postoperative development of new cerebral ischemic lesions caused by intraoperative microemboli. Neurosurgery 2011. 10.1227/NEU.0b013e318214abf6.

MacDonald DB, Dong C, Quatrale R, Sala F, Skinner S, Soto F, et al. Recommendations of the International Society of Intraoperative Neurophysiology for intraoperative somatosensory evoked potentials. Clin Neurophysiol 2019;130:161–79. 10.1016/j.clinph.2018.10.008.

MacDonald DB, Skinner S, Shils J, Yingling C. Intraoperative motor evoked potential monitoring – A position statement by the American Society of Neurophysiological Monitoring. Clin Neurophysiol 2013;124:2291–316. 10.1016/j.clinph.2013.07.025.

Muramoto A, Imagama S, Ito Z, Wakao N, Ando K, Tauchi R, et al. The cutoff amplitude of transcranial motor-evoked potentials for predicting postoperative motor deficits in thoracic spine surgery. Spine (Phila Pa 1976) 2013;38:E21–7. 10.1097/BRS.0b013e3182796b15.

Neuloh G, Pechstein U, Cedzich C, Schramm J. Motor evoked potential monitoring with supratentorial surgery. Neurosurgery 2004;54:1061–70; discussion 1070-2. 10.1227/01.neu.0000119326.15032.00.

Neuloh G, Simon M, Schramm J. Stroke prevention during surgery for deep-seated gliomas. Neurophysiol Clin Neurophysiol 2007;37:383–9. 10.1016/j.neucli.2007.09.002.

Noel-Storr AH, McCleery JM, Richard E, Ritchie CW, Flicker L, Cullum SJ, et al. Reporting standards for studies of diagnostic test accuracy in dementia: The STARDdem Initiative. Neurology 2014;83:364–73. 10.1212/WNL.0000000000000621.

Nuwer MR, Dawson EG, Carlson LG, Kanim LEA, Sherman JE. Somatosensory evoked potential spinal cord monitoring reduces neurologic deficits after scoliosis surgery: results of a large multicenter survey. Electroencephalogr Clin Neurophysiol Evoked Potentials 1995. 10.1016/0013-4694(94)00235-D.

Nwachuku EL, Balzer JR, Yabes JG, Habeych ME, Crammond DJ, Thirumala PD. Diagnostic value of somatosensory evoked potential changes during carotid endarterectomy: A systematic review and meta-analysis. JAMA Neurol 2015;72. 10.1001/jamaneurol.2014.3071.

Pan S-Y, Holdefer RN, Wu H-L, Li C-R, Guo L. The predictive value of intraoperative facial motor evoked potentials in cerebellopontine angle tumor surgery. Clin Neurophysiol 2024;166:176–90. 10.1016/j.clinph.2024.07.021.

Radtke RA, Erwin CW, Wilkins RH. Intraoperative brainstem auditory evoked potentials: Significant decrease in postoperative morbidity. Neurology 1989;39:187–187. 10.1212/WNL.39.2.187.

Reddy RP, Brahme IS, Karnati T, Balzer JR, Crammond DJ, Anetakis KM, et al. Diagnostic value of somatosensory evoked potential changes during carotid endarterectomy for 30-day perioperative stroke. Clin Neurophysiol 2018;129. 10.1016/j.clinph.2018.05.018.

Sala F, Manganotti P, Tramontano V, Bricolo A, Gerosa M. Monitoring of motor pathways during brain stem surgery: what we have achieved and what we still miss? Neurophysiol Clin 2007;37:399–406. 10.1016/j.neucli.2007.09.013.

Sala F, Palandri G, Basso E, Lanteri P, Deletis V, Faccioli F, et al. Motor evoked potential monitoring improves outcome after surgery for intramedullary spinal cord tumors: a historical control study. Neurosurgery 2006;58:1129–43; discussion 1129-43. 10.1227/01.NEU.0000215948.97195.58.

Sanai N, Mirzadeh Z, Berger MS. Functional Outcome after Language Mapping for Glioma Resection. N Engl J Med 2008;358:18–27. 10.1056/NEJMoa067819.

See RB, Awosika OO, Cambria RP, Conrad MF, Lancaster RT, Patel VI, et al. Extended Motor Evoked Potentials Monitoring Helps Prevent Delayed Paraplegia After Aortic Surgery. Ann Neurol 2016;79:636–45. 10.1002/ana.24610.

Seidel K, Beck J, Stieglitz L, Schucht P, Raabe A. The warning-sign hierarchy between quantitative subcortical motor mapping and continuous motor evoked potential monitoring during resection of supratentorial brain tumors. J Neurosurg 2013;118:287–96. 10.3171/2012.10.JNS12895.

Seidel K, Szelényi A, Bello L. Intraoperative mapping and monitoring during brain tumor surgeries. In: Deletis V, Shils JL, Sala F, Seidel K, editors. Neurophysiol. Neurosurg. A Mod. Approach. 2nd ed., Academic Press; 2022, p. 133–49. 10.1016/B978-0-12-819826-1.00013-2.

Seidel K, Wagner N, Wermelinger J, Abut PA, Branca M, Sivanrupan S, et al. Motor evoked potential monitoring and continuous dynamic mapping: Warning criteria during surgery on motor eloquent intra-axial brain tumors. Clin Neurophysiol 2025;178:2110979. 10.1016/j.clinph.2025.2110979.

Skinner S, Holdefer R, McAuliffe JJ, Sala F. Medical Error Avoidance in Intraoperative Neurophysiological Monitoring: The Communication Imperative. J Clin Neurophysiol 2017;34:477–83. 10.1097/WNP.0000000000000419.

Skinner S, Sala F. Communication and collaboration in spine neuromonitoring: time to expect more, a lot more, from the neurophysiologists. J Neurosurg Spine 2017;27:1–6. 10.3171/2016.12.SPINE161212.

Skinner SA, Holdefer RN. Intraoperative neuromonitoring alerts that reverse with intervention: treatment paradox and what to do about it. J Clin Neurophysiol 2014;31:118–26. 10.1097/WNP.0000000000000030.

Szelényi A, Bello L, Duffau H, Fava E, Feigl GC, Galanda M, et al. Intraoperative electrical stimulation in awake craniotomy: methodological aspects of current practice. Neurosurg Focus 2010;28:E7. 10.3171/2009.12.FOCUS09237.

Szelényi A, Langer D, Kothbauer K, De Camargo AB, Flamm ES, Deletis V. Monitoring of muscle motor evoked potentials during cerebral aneurysm surgery: intraoperative changes and postoperative outcome. J Neurosurg 2006;105:675–81. 10.3171/jns.2006.105.5.675.

Thirumala PD, Carnovale G, Habeych ME, Crammond DJ, Balzer JR. Diagnostic accuracy of brainstem auditory evoked potentials during microvascular decompression. Neurology 2014;83. 10.1212/WNL.0000000000000961.

Toleikis JR, Pace C, Jahangiri FR, Hemmer LB, Toleikis SC. Intraoperative somatosensory evoked potential (SEP) monitoring: an updated position statement by the American Society of Neurophysiological Monitoring. J Clin Monit Comput 2024;38:1003–42. 10.1007/s10877-024-01201-x.

Whiting P, Rutjes AWS, Reitsma JB, Glas AS, Bossuyt PMM, Kleijnen J. Sources of variation and bias in studies of diagnostic accuracy: a systematic review. Ann Intern Med 2004;140:189–202. 10.7326/0003-4819-140-3-200402030-00010.

Whiting PF, Rutjes AWS, Westwood ME, Mallett S, Deeks JJ, Reitsma JB, et al. QUADAS-2: a revised tool for the quality assessment of diagnostic accuracy studies. Ann Intern Med 2011;155:529–36. 10.7326/0003-4819-155-8-201110180-00009.

Wilent WB, Tesdahl EA, Harrop JS, Welch WC, Cannestra AF, Poelstra KA, et al. Utility of motor evoked potentials to diagnose and reduce lower extremity motor nerve root injuries during 4,386 extradural posterior lumbosacral spine procedures. Spine J 2020;20:191–8. 10.1016/j.spinee.2019.08.013.

De Witt Hamer PC, Robles SG, Zwinderman AH, Duffau H, Berger MS. Impact of Intraoperative Stimulation Brain Mapping on Glioma Surgery Outcome: A Meta-Analysis. J Clin Oncol 2012;30:2559–65. 10.1200/JCO.2011.38.4818.

