## Supplementary Table 1 for "Standards for Reporting Diagnostic Accuracy Studies Using Intraoperative Neurophysiological Monitoring"

| Supplementary Table 1: IONM-specific commentary and examples of adequate reporting with citation. | | | |
| --- | --- | --- | --- |
| Section,  and  Item | Item | IONM specific commentary | IONM-specific examples with citation. |
| TITLE OR ABSTRACT | | | |
| 1 | Identify the report as a diagnostic accuracy study using at least one accuracy measure. (such as but not limited to sensitivity, specificity, predictive values, or AUC) | When possible, titles should explicitly include terms such as intraoperative and diagnostic accuracy, or refer to predictive performance (e.g., sensitivity, specificity). Including “intraoperative” improves discoverability in systematic reviews. In some cases, especially when describing new monitoring methods, full diagnostic terminology may be difficult to incorporate, but emphasis on predictive metrics is recommended. | Clearly identifies a diagnostic-accuracy study in IONM and names an accuracy metric, enabling rapid retrieval and indexing: Diagnostic accuracy of somatosensory evoked potential and electroencephalography during carotid endarterectomy. Results: Maximum sensitivity was obtained with multimodality monitoring with an IONM change in either EEG or SSEP of 50.00 (95% CI, 30.66-69.34). The specificity of simultaneous EEG and SSEP changes was 93.95 (95% CI, 92.28-95.35%). Maximum area under ROC curve obtained for IONM change in either EEG or SSEP was 0.660 (95% CI, 0.547-0.773, p-value 0.004).(Thirumala et al., 2016) The diagnostic accuracy of neuromonitoring for detecting postoperative bowel and bladder dysfunction in spinal oncology surgery: a case series, Sacral reflexes exhibited a specificity and sensitivity of 100% and 73.33%, respectively, with a positive predictive value (PPV) of 100% and a negative predictive value (NPV) of 89.19% for predicting immediate post-operative BBD. At discharge and the 1-month follow-up, the specificity and PPV remained at 100%. However, the sensitivity dropped to 64.71%, and the NPV decreased to 83.78%.(Silverstein et al., 2024) |
| ABSTRACT | | | |
| 2 | Structured summary of study design, methods, results, and conclusions (for specific guidance, see STARD for Abstracts) | The abstract should clearly and concisely state the aim or hypothesis. The methods section must specify study type (e.g., prospective, retrospective, observational), number of patients, and statistical analyses performed. Results should report key findings, including sample size and main outcomes (e.g., odds ratios, confidence intervals, p-values, predictive values). Conclusions should interpret the clinical relevance for IONM practice, indicate whether further studies are needed, and summarize the significance of the findings. | Provides a structured abstract with design, sample, methods, key accuracy estimates, and conclusions in one place: Objective: To develop and evaluate machine learning (ML) approaches for muscle identification using intraoperative motor evoked potentials (MEPs), and to compare their performance to human experts.(Boaro et al., 2024) Methods: The authors prospectively studied 69 patients who underwent tumor surgery adjacent to the CST (< 1 cm using diffusion tensor imaging and fiber tracking) with simultaneous subcortical monopolar motor mapping (short train, interstimulus interval 4 msec, pulse duration 500 μsec) and a new acoustic motor evoked potential alarm. Continuous (temporal coverage) and dynamic (spatial coverage) mapping was technically realized by integrating the mapping probe at the tip of a new suction device, with the concept that this device will be in contact with the tissue where the resection is performed. Motor function was assessed 1 day after surgery, at discharge, and at 3 months.(Raabe et al., 2014) Results: A total of 563 patients underwent aortic arch reconstruction with HCA and IONM. Of these, 119 (21.1%) patients had an IONM change, whereas 444 (78.9%) did not. Patients with IONM changes had increased operative mortality (22.7% vs4.3%) and increased early ANE (10.9% vs 2.9%). In multivariable analysis, SSEP changes were correlated with early ANE (odds ratio [OR], 4.68; 95% confidence interval [CI], 1.51-14.56; P ¼ .008), whereas EEG changes were not (P ¼ .532). Permanent SSEP changes were correlated with early ANE (OR, 4.56; 95%CI, 1.51-13.77; P ¼ .007), whereas temperature-related SSEP changes were not (P ¼ .997). Finally, any IONM change (either SSEP or EEG) was correlated with operative mortality (OR, 5.82; 95% CI, 2.72-12.49; P<.001).(Sultan et al., 2020) Conclusions: While TcMEPs and TNSEPs were found to be reliable intraoperative neurophysiological monitoring parameters during tethered cord syndrome surgery, PAR had low sensitivity and positive predictive value probably because the reflex is not directly related to bladder function and because its multisynaptic pathway may be sensitive to anesthetics. New onset muscle weakness and sensory deficits were related to postoperative changes in TcMEPs and TNSEPs, whereas changes in PAR did not predict bladder/urinary impairment. Urinary deficits may be predicted and prevented with other neurophysiological techniques, such as the bladder-anal reflex.(Squintani et al., 2025) |
| INTRODUCTION | | | |
| 3 | Scientific and clinical background, including the intended use and clinical role of the index test | A description of the scientific and clinical background is important, starting with a broad description of the field, of current knowledge and then identification of the knowledge gap. Information about the reference test, and its shortcomings, is important. Information about the rationale for a different or newer test (index test) should be given, and where relevant the knowledge gap. When the reference test is clinical outcome, the authors should clearly state which outcome parameter is used; and ideally explain why there is no intraoperative reference test. | Concisely links clinical need, index test role, and reference standard limitations to a focused research gap: Scientific and clinical background: "Intraoperative neurophysiological monitoring (IONM) has become the gold standard for spinal cord monitoring during spinal surgery. Transcranial electrical stimulation motor-evoked potentials (Tc-MEPs) are used to monitor the motor pathways, and somatosensory-evoked potentials (SSEPs) are used to monitor the sensory pathways. Tc-MEPs can be recorded directly in the epi- or subdural space over the spinal cord using direct-waves (D-waves) or from the muscles (mTc-MEPs). mTc-MEPs can be recorded using either extramuscular (surface or subcutaneous needle) or intramuscular (needle or hook wire) electrodes. When mTc-MEPs are recorded with extra muscular electrodes, volume conduction significantly affects the recorded potentials by influencing the waveform morphology, decreasing the measured amplitudes, and changing the frequency content of the signals. Electromyography (EMG) and intraoperative motor root studies have shown that spontaneous discharges, e.g., myotonic discharges, fibrillations, and positives spikes, cannot be adequately registered using surface electrodes. Moreover, Skinner et al.v concluded that EMG monitoring during myelopathy surgery should not be performed using surface electrodes. It was stated that intramuscular electrodes are preferred for lower motor neuron function monitoring, since near field recordings are required. An example of a good description of the knowledge gap is: "It is however not known whether mTc-MEP recordings from surface electrodes are of equivalent utility to mTc-MEPs recorded from subcutaneous needles for detection of mTc-MEP warnings during spinal cord monitoring. "(Dulfer et al., 2023) |
| 4 | Study objectives and hypotheses | Clearly state the primary objective of the study, such as assessing the diagnostic accuracy of an IONM modality (e.g., SSEP, MEP, EEG) to predict perioperative neurological deficits. Hypotheses should reflect expected performance measures like sensitivity, specificity, or predictive values. Pre-specifying primary and secondary hypotheses strengthens the methodological rigor and reduces bias in interpretation.  IONM-specific considerations: In studies involving IONM, objectives may include determining whether specific intraoperative changes predict postoperative deficits, or whether the absence of changes reliably indicates neurological preservation. Hypotheses should address the relationship between IONM signal changes (index test) and the occurrence of postoperative outcomes (reference standard), while accounting for the potential influence of intraoperative interventions. | States a testable hypothesis and primary objective aligned with an accuracy endpoint: “ Objective: To evaluate the ability of intraoperative neurophysiologic monitoring (IONM) during aortic arch reconstruction with hypothermic circulatory arrest (HCA) to predict early (<48 hours) adverse neurologic events (ANE; stroke or transient ischemic attack) and operative mortality.(Sultan et al., 2020)  Objective was to evaluate the feasibility of IONM time CCEPs in a cohort of glioma patients undergoing minimally invasive surgery in an asleep-awake-asleep setting. Compared the quality of CCEPs obtained during the awake and asleep phases. Also compared CCEP properties according to tumor location, histopathology, sex, and aphasia. Hypothesis was that CCEP may be able to be used reliably in both the awake and asleep settings for language mapping.  Concluded that CCEP recordings are feasible during minimally invasive surgery and might be surrogate markers for altered connectivity of the language tracts.(Seidel et al., 2024)” |
| METHODS | | | |
| Study design | | | |
| 5 | Specify whether the study was prospective or retrospective. | This is certainly important for IONM studies. It is particularly needed when interpreting the adequacy of the study design and interpreting the study results. If prospective, then one can determine whether important confounds were avoided by perusing the eligibility criteria and methods; and checking that an adequate sample size calculation was performed. For prospectively collected data, it is important to know whether how the results of the index or reference standard were interpreted, and whether action was taken (i.e. more information needed later in the methods). If the data were collected retrospectively, then subsequent descriptions in the methods and results of attempts to adjust for confounders are important. | Explicitly declares prospective/retrospective design and timing of test and reference standard: This study was approved by the institutional review board of the Seoul National University Hospital (IRB approval no., H-1707-003-864) and the Ethics Committee exempted the study for obtaining patient informed consent as this was a minimal risk study. We retrospectively reviewed 317 cases from a single-institution prospective cohort of patients who underwent spine surgery using multimodal IONM at the department of orthopedic surgery between January 2013 and May 2017.(Wi et al., 2020)  "In this analysis, we performed a prospective multicenter cohort study in 16 Japanese spinal centers with the WG of the JSSR from April 1, 2017 to March 31, 2018 (Figure 1). This assignment was approved by each institutional review board in the participating centers."(Takahashi et al., 2021) |
| Participants | | | |
| 6 | Eligibility criteria | Clear eligibility criteria—age, sex, diagnosis, comorbidities,—are essential in STARD-IONM to reduce bias, ensure reproducibility, and support valid comparisons and clinical applicability. | Specifies inclusion/exclusion (age, diagnosis, surgical context), supporting reproducibility and applicability: This is a reasonably clear description of eligibility criteria: " In order to be eligible to participate in this study, patients must meet all the following criteria: – ≥12 years. Demonstrated spinal pathology for which surgery with the use of mTc-MEP monitoring has been planned. Signed and dated informed consent document prior to any study-related procedures. A potential subject who meets any of the following criteria will be excluded from participation in this study: Patient refusal. Existing motor weakness in the left or right tibialis anterior, gastrocnemius, or abductor hallucis muscles. History of epilepsy. Contra-indications to IONM such as presence of a pacemaker or implantable cardioverter-defibrillator. Patients with history of stroke or cranial lesions, increased intracranial pressure, heart failure and longstanding hypertension."(Dulfer et al., 2021) |
| 7 | Describe how potentially eligible participants were identified. | It is important to define eligibility criteria and describe how individuals were selected to ensure the study hypothesis can be appropriately investigated. Authors should specify how and where eligible subjects were identified, and report detailed inclusion and exclusion criteria to allow reproducibility. | Describes how candidates were identified (symptoms/registry/prior tests) and shows the screening pathway: The authors analyzed data obtained in a series of 100 consecutive patients with intrinsic brain tumors (n = 76), metastatic lesions (n = 17), and cavernous malformations (n = 7) close (≤ 10 mm) to the CST, as shown on preoperative DTI fiber tracking. The tumor surgeries were performed from March 2009 until October 2011. We included only patients in whom the craniotomy and tumor approach allowed placement of a strip electrode on the precentral gyrus for DCS monitoring. (Seidel et al., 2013) |
| 8 | Specify where and when potentially eligible participants were identified. | Report where and when eligible participants were identified, including details such as patient pool, clinical setting, age range, condition, and team experience. This contextualizes study applicability, minimizes selection bias, and improves reproducibility. | Reports setting, location(s), and calendar dates, allowing assessment of external validity.: “All patients included in our analysis had been referred to our department (Neurosurgical Clinic, Clinic of the University of Munich [LMU], Germany) in the period from January 2000 to December 2013 for the resection of spinal cord hemangioblastoma. After obtaining study approval from the IRB of the Clinic of the University of Munich (LMU), Germany, we identified and retrospectively reviewed medical records and radiological studies for 24 patients from our clinic database. (Siller et al., 2017)” |
| 9 | Whether participants formed a consecutive, random or convenience series | Random or convenience sampling can introduce selection bias. Consecutive enrollment of subjects, whether in prospective or retrospective studies, helps minimize bias and improves study validity. | States enrollment method (consecutive/random/convenience) and justifies any deviations, clarifying selection bias: "A single center retrospective analysis of consecutive pediatric patients requiring cervical spine fixation and/or decompression between March 2015 – February 2022 was performed. All patients included in our analysis had been referred to our department (Neurosurgical Clinic, Clinic of the University of Munich [LMU], Germany) in the period from January 2000 to December 2013 for the resection of spinal cord hemangioblastoma. After obtaining study approval from the IRB of the Clinic of the University of Munich (LMU), Germany, we identified and retrospectively reviewed medical records and radiological studies for 24 patients from our clinic database. "(McDevitt et al., 2022) "We evaluated the prospectively collected neuromonitoring data of 354 consecutive operated cases of adolescent idiopathic scoliosis, from an ongoing prospective study, by independent observer (2004-2008)."(Kundnani et al., 2010) |
| Test methods | | | |
| 10 | Describe the index test with technical specifications sufficient for replication. | Describe the index test in sufficient detail to permit replication, including modality, stimulation and recording parameters, equipment, acquisition timing, baseline definition, alert criteria, and any patient- or procedure-specific adaptations. Report anesthesia and physiology variables that may affect interpretation, including anesthetic technique, neuromuscular blockade, at baseline and at alerts.  Authors should describe the index test methodology with sufficient technical detail to permit replication. This includes specifying the stimulation and recording parameters (e.g., stimulus type, intensity, frequency, recording montage, filter settings), equipment used (manufacturer and model if relevant), and any modifications or special techniques applied during data acquisition. The description should reflect real intraoperative conditions and clarify any patient- or procedure-specific adaptations made to standard protocols. Include anesthesia-relevant factors affecting IONM (technique TIVA/volatile/mixed and NMB level before MEPs, MAP/ at baseline and at alerts). | Details stimulation/recording parameters, baseline timing, averaging/filters, modality sequence, and any intra-op parameter changes—making replication feasible: Motor evoked potentials: Stimulating electrodes were placed according to the international 10/20 system of electrode placement. Corkscrew needle electrodes were placed in the C1, C2, C3, and C4 locations. Additionally, electrodes were placed in M1, M2, M3, and M4; these locations are 1 cm anterior to the corresponding C electrodes. The M locations were used because these sites have been reported to be closer to the motor cortex than the C sites. A constant voltage stimulator was used. The TES pulse width was 75 microseconds, interstimulus interval was 2.1 milliseconds, and a single train of 5 to 7 pulses was used. Stimulation intensity was held constant after finding the threshold for the foot MEP using the control stimulation montage.(Schwartz et al., 2022) Blink reflex: Stimulation of the supraorbital nerve of either side of the face was performed on the first 5 patients using a pair of electroencephalographic needle electrode (disposable twisted pair subdermal needle electrode; Nicolet BioMedical, Inc., Madison, Wisconsin) inserted subcutaneously over the supraorbital nerve, whereas a surface electrode (disposable disk electrodes; Viasys Healthcare, Madison, Wisconsin) was used on the remaining patients. One to seven rectangular constant-current stimuli with an interstimulus interval (ISI) of 2 ms, intensity of 20–40 mA, and train repetition rate of 0.4 HZ were used. A double train of stimuli was used on 5 patients, with the intertrain interval ranging from 20 to 40 ms. Recording was done with needle electrodes identical to those used for stimulation. The electrodes were inserted in the low lateral part of the orbicularis oculi muscle ipsilateral to the stimulating side after identifying muscle twitches evoked by stimulating facial nerve branches. Recording was performed after averaging two single sweeps. After the first sweep, the polarity of the stimulating electrode was reversed to avoid a large stimulus artifact. Recordings were made by using a 50-ms epoch and bandpass digital filters of 70 and 1219 HZ. Recording of the BR was attempted after intubation, in the middle of surgery, and after starting skin closure.(Deletis et al., 2009) |
| 11 | Describe the reference standard(s). | Describe the reference standard in sufficient detail to permit replication, including the neurologic or functional domain assessed, timing of assessment, procedures used, thresholds/categories, assessor qualifications, and whether assessors were blinded to index-test results. When postoperative outcomes are used, specify the time point and whether they were compared with the final IONM signal recorded near the end of surgery. | Defines the reference standard with procedures, timing, assessors, and thresholds, enabling valid comparison: "The primary outcome of interest was the presence of postoperative delirium (POD) during the patient's ICU stay after surgery. Delirium was assessed multiple times during the patient's ICU stay using the Intensive Care Delirium Screening Checklist (ICDSC) scores. The ICDSC is a scoring system that ranges from 0 to 8 and is used to assess the presence of delirium. Throughout a 12-hour shift, the registered nurse's observations of the patient are used to calculate the ICDSC score at the end of the shift. Patients with a single ICDSC score ≥4 are considered to have delirium." (Al-Qudah et al., 2024) "As a routine part of workup and follow-up, all patients underwent formal neurological evaluation immediately before the procedure, postoperatively prior to discharge, and at 3– to 6-month clinical follow-up. Clinical status was established according to a modified version of the McCormick grading scale, which is a functional scale designed to account for both motor and sensory functions"(Korn et al., 2015) " "As for hearing function, pure tone audiometry was performed on both sides 2–4 days before and 8–10 days after surgery, and pure tone average (PTA) was used to define hearing impairment after MVD. In audiometry, PTA represents the average hearing thresholds for 500, 1000, 2000, and 4000 Hz. High-frequency hearing loss (HFHL) was also examined, defined as an increase of more than 10 dB in the hearing threshold at either 4000 Hz or 8000 Hz"(Parthasarathy D. Thirumala et al., 2014) "perioperative stroke (i.e., any confirmed neurologic deficit of abrupt onset noted in medical record caused by a disturbance in blood supply to the brain within the first 24 hours after the procedure, including ophthalmic and retinal strokes) or transient ischemic attack (i.e., any neurologic deficit of abrupt onset noted in medical record caused by a disturbance in blood supply to the brain within the first 24 hours after the procedure and lasted less than 24 hours). In this investigation, only patients with neurologic complaints or overt signs were assessed postoperatively for stroke."(Thirumala et al., 2021)  Rationale for choosing the reference standard (if alternatives exist)  Justifies the chosen reference standard among alternatives and discusses its limitations: "Ohy-Maldaun Fast Track Cognitive Test (OMFTCT). OMFTCT is the second step of our ongoing Language Tracking Project and focuses on assessing production and comprehension at main linguistic domains such as phonology, semantics, and morphosyntax. OMFTCT domains and tasks were chosen based on Wechsler Intelligence Scales, Raven’s Progressive Matrices, Rey’s Complex Figures, Benton’s Visual Retention Test and the Token Test. A systematic review identified the most used tests in patients with glioma, aiming to detect impairments in memory, attention, and executive functions. The neurological state of each patient was evaluated before surgery, immediately after surgery, at 1, 6 and 12 months later and then once per year thereafter. The clinical status was established according to a modified version of the McCormick grading scale, which is a functional scale designed to account for both motor and sensory functions.(Ohy et al., 2024) As the McCormick grading scale is hard to differentiate between radiculopathy and myelopathy, in our study we classified patients with myelopathy."(Rho et al., 2016) |
| 12a | Define and justify index-test positivity cut-offs/result categories; distinguish prespecified vs exploratory. | Define the index-test positivity threshold(s) or result categories and justify their use, including whether thresholds were pre-specified or data-derived.  The interpretation of the index test depends on how positivity is defined. Researchers must specify the exact threshold, cutoff value, or result category used to classify the index test as positive or negative. This includes, for example, amplitude changes, signal loss percentages, or categorical outputs in tests like SSEP, Brainstem auditory evoked potentials, EEG, or MEPs. The rationale for choosing this threshold should be provided, based on physiological reasoning, clinical guidelines, or prior studies. It must also be clearly stated whether the definition was determined before data collection (pre-specified) or derived from the study data itself (exploratory). Pre-specified cut-offs enhance reproducibility and reduce bias, while exploratory cut-offs should be interpreted cautiously and validated in future studies. State how alert criteria account for anesthetic depth, neuromuscular blockade, and physiologic state; specify rules for near-threshold fluctuations. | Prespecifies alert/positivity criteria (magnitude and persistence), and explains handling of near-threshold fluctuations: Motor evoked potential: The published internal practice guidelines for alerts used by the IONM teams in this study stated, “Alarm criteria for transcranial electric MEPs can vary, depending on size and variability of responses, abruptness of change as well as other factors. In practice, 75-80% attenuation of MEP amplitude during spine surgery warrants an alert, although changes that are smaller than this may also be significant.” No established criterion for spinal nerve root monitoring has been established. This IONM group has historically employed a 50% amplitude reduction as a part of the criterion for C5 nerve root dysfunction. Thus, the change in amplitude that prompted an MEP alert for putative nerve root injuries in this dataset was at the clinical discretion of the IONM team, after weighing multiple variables, but it was often a 50% attenuation that prompted an alert. (Wilent et al., 2020)  Multimodal monitoring: Regarding neurophysiologic changes during the period of ICA cross-clamping, tSSEP and tcMEP were compared with mSSEP parameters. A change in EP monitoring was “significant” when the following EP alterations occurred: (1) a reproducible decrement (>50%) of ipsilateral elicited mSSEP N20/P25 amplitudes; (2) a reproducible decrement (>50%) of ipsilateral elicited tSSEP P40/N50 amplitudes; and (3) the loss of contralateral recorded tcMEP responses (all-or-none interpretation). The baseline was set within the last measurements of SSEPs and tcMEP amplitudes before ICA cross-clamping. (Malcharek et al., 2015)  In the evaluation of intraoperative facial muscle corticobulbar motor evoked potentials (FMcoMEPs), several diagnostic test definitions and thresholds are used to guide surgical decision-making. The Bilateral Monitoring Technique (BilatMT) compares the difference in stimulation threshold increases between the ipsilateral and contralateral facial muscles from baseline to the end of resection. In the optimistic approach, a test is considered negative if the difference remains under 20% in all ipsilateral muscles, while in the traditional approach, the threshold applies to at least one facial muscle. The Unilateral Monitoring Technique (UnilatMT) evaluates only the ipsilateral side, with the optimistic approach defining a negative test as requiring less than a 20-mA increase in stimulation threshold across all ipsilateral muscles, and the traditional approach applying the same threshold to just one muscle. Intraoperative warnings are triggered by physiological changes such as prolonged A-train activity, a greater than 50% amplitude reduction necessitating increased stimulation intensity, transient FMcoMEP loss, or permanent FMcoMEP loss. The presence of these findings prompts an intraoperative alert to the surgeon, whereas their absence indicates no warning was issued. These definitions help standardize IONM responses and support timely intervention to prevent postoperative deficits."(Greve et al., 2021) |
| 12b | Define and justify reference standard positivity cut-offs and result categories; distinguish prespecified vs exploratory. | Define the reference-standard positivity threshold(s) or result categories and justify their use, including timing of outcome assessment, diagnostic confirmation methods, and whether categories were pre-specified or exploratory. If assessor judgment was involved, describe steps taken to reduce variability and bias.  The reference standard is the benchmark used to judge the accuracy of the index test. Researchers must clearly define what constitutes a positive result, such as the presence of a clinical condition, the timing of evaluation, and use of diagnostic test confirmation (audiogram for hearing, MRI for stroke) or specific clinical outcome (e.g. delirium). The criteria should be explicitly described and supported by clinical rationale or standard definitions (e.g NIH stroke scale, MRC grading scale, ICDSC scale for delirium). Importantly, the report must state whether these definitions were established before the study began (pre-specified) or determined after examining the data (exploratory). When outcome definitions are based on clinical judgment or follow-up, the risk of variability and bias increases—particularly if the assessor is influenced by index test results. Transparent reporting allows readers to assess the reliability and applicability of the study’s accuracy estimates. | Prespecifies reference-standard categories/time windows and distinguishes prespecified vs exploratory cut-offs: “Facial nerve outcomes: "To perform risk stratification, the outcome measure (facial muscle function) had to be dichotomized. We compared two forms of dichotomization: The first was to dichotomize between “no deterioration in House–Brackmann score” and “any deterioration in House–Brackmann score”. The second was to dichotomize between “no/mild deterioration” and “relevant deterioration”. No/mild deterioration was defined as either no deterioration or an increase in House–Brackmann score without exceeding an absolute score of III (complete eye closure preserved). Relevant deterioration was defined as deterioration of the House–Brackmann score with an absolute value of IV or higher (including incomplete eye closure). This second form of dichotomization was introduced because incomplete eye closure carries a high risk of secondary complications and because transitions of the House–Brackmann score from one point to the next have been shown to have high interobserver variability. In contrast, incomplete eye closure represents a clearer clinical parameter for dichotomization than small changes in the House–Brackmann score. Despite the theoretical advantages explained above, we used ROC curve analysis to determine whether this new form of outcome dichotomization is a valid alternative to evaluating stepwise changes in the House–Brackmann score.(Greve et al., 2021) Audiologic investigations and HL criteria. We considered tone audiometry consisting of PTA scores and SDS to be the indicator of auditory function.13,14 Postoperative HL status was assessed using the 1995 AAO-HNS classification system.8 Class B referred to useful or serviceable HL and Class C/D was nonservice able HL that was not amenable to hearing aids. We performed an otoneurologic examination on all patients preoperatively (median 1 day, range 1–49 days) and postoperatively (median 7 days, range 1–90 days). This consisted of an audiogram with measurement of pure tone thresholds (air and bone conduction for octave frequencies 250–8,000 Hz) as previously described. All audiograms were performed at the Eye and Ear Institute, University of Presbyterian Hospital, by an audiologist.(Parthasarathy D. Thirumala et al., 2014)” |
| 13a | Specify whether clinical information and reference standard results were available to index-test assessors. | State whether performers/readers of the index test had access to clinical information, operative context, or reference-standard results, and describe how this may have influenced interpretation.  In diagnostic accuracy studies, especially those involving tests requiring subjective interpretation—such as imaging, electrophysiological monitoring, or clinician-judged outcomes—it is critical to report whether the interpreters of each test had access to other information that could influence their judgment. Item 13a requires reporting whether clinical details (e.g., symptoms, surgical findings) or reference standard outcomes (e.g., postoperative deficits) were known to the individuals performing or reading the index test. If such information was available, their interpretation might be biased toward the expected result, leading to inflated diagnostic accuracy. | States whether index-test performers/readers had access to clinical or reference results, clarifying potential bias: “Index Test: The randomization of the study (1:1) was performed by the narcotic drug administrator in the Department of Anesthesia, using internet-based randomization software. The morning of the surgery blinded halogenated anesthetics (visually identical plastic bottles of 100 ml) were delivered to the anesthesiologist in charge of the patient. Study anesthetics were only identified by the assigned patient number. Intraoperative electrophysiological data were analyzed and recorded by one neuroelectrophysiological specialist. Importantly, the neuroelectrophysiological specialist remained blinded to the anesthetic allocation and the inspiratory concentration when collecting data.(Xiang et al., 2021)  IONM was conducted using the Cascade® IONM system (Cadwell Industries) by a skilled technician under the direct supervision of a professional physiatrist blinded to the group allocation of patients. Intraoperative SEP waves were acquired continuously throughout the surgical procedure, following a protocol like that used for preopSEPs. The initial waveform was acquired immediately after completion of the IONM setup that occurred after anesthesia induction. (Kim et al., 2025) |
| 13b | Specify whether index-test results were available to reference-standard assessors. | Item 13b addresses whether those assessing the reference standard (e.g., determining postoperative neurological outcome) had access to the index test results. If so, their judgment might be influenced by knowledge of IONM alerts or lack thereof, artificially increasing agreement between tests. This is particularly relevant in IONM studies where surgical teams might anticipate or dismiss subtle deficits based on intraoperative monitoring trends.  This reporting item does not prescribe whether blinding is necessary—it simply requires that the presence or absence of blinding be transparently described. This enables readers and reviewers to understand the context of test interpretation and judge the potential risk of bias in accuracy estimates. If full blinding was not feasible, explain what information was available, and why, so the study’s generalizability to real-world practice can be assessed. | Reference Standard: A new neurologic deficit was judged by a surgeon who was blinded to the neuromonitoring results. The overall rate of a new  neurologic deficit was 20% (n = 84 of 423). The overall rate of an intervention among all patients was 10% (n = 42 of 423). The rate of a new neurologic deficit among those patients who received an intervention was 4.7% and among those who did not receive an intervention was 15.1%. The authors reported that  those patients who did not receive an intervention may have had alerts that did not warrant an intervention. The authors reported that 5.2% of the patient’s benefited from monitoring based on those patients who awoke without neurologic deficits (n = 22 of 423) when there was a positive intraoperative alert.(Wiedemayer et al., 2002)  Medical records for all 508 patients were reviewed to determine if any new neurologic deficits developed when the patient woke up from anesthesia. The medical records were reviewed independently without knowledge of the intraoperative SSEP changes in a blinded fashion. Any new postoperative motor/sensory deficits or bowel/bladder changes were iatrogenic intraoperative injuries. Similarly, the intraoperative SSEPs for all 508 patients were reviewed independently without knowledge of the postoperative neurologic outcome in a blinded fashion.(Khan et al., 2006)  Partial Blinding: From study onset, the results of all intraoperative testing were withheld from the surgical team; that is, the team was blinded to test results. Beginning with the third patient tested, we added a provision to provide feedback to the surgical team—to break the blind—but only if testing indicated a high probability that the stimulating probe was in physical contact with, or in close proximity to, the dura mater of the spinal cord; a screw placed under these circumstances could cause either immediate or delayed injury to the spinal cord, so this risk needed to be avoided. We refer to this portion of the study in which only one type of feedback (that is, break the blind) was used as Phase 1. In the final year of the study, we entered Phase 2, during which we started providing feedback to the surgical team on a regular basis.(Calancie et al., 2014)” |
| Analysis | | | |
| 14 | Methods for estimating or comparing measures of diagnostic accuracy | Describe the statistical methods used to estimate or compare diagnostic accuracy, including sensitivity, specificity, predictive values, likelihood ratios, odds ratios, AUC, and confidence intervals. State how variability was assessed, how comparisons between IONM modalities were made, and how ND/PR/RD/ID categories were handled in the analysis.  Authors must describe in detail how diagnostic performance was measured. This includes calculating sensitivity, specificity, predictive values (PPV, NPV), likelihood ratios, diagnostic odds ratios, and area under the ROC curve (AUC). Clearly specify the type of each variable analyzed—continuous, ordinal, categorical, or binary—and for discrete variables, report all possible values or ranges. State which statistical methods were used to obtain p-values and confidence intervals for diagnostic measures (e.g., chi‑square or Fisher’s tests, logistic regression, bootstrap methods for CIs). When comparing multiple IONM modalities, describe comparative tests used (e.g., McNemar’s test for paired proportions, DeLong’s test for AUC comparisons). Additionally, accuracy analyses should account for reversible IONM changes using extended contingency tables (e.g., 3×2 format as outlined by Holdefer et al.), and regression models (logistic or ordinal) should be named and their use justified when adjusting for covariates. Explicit reporting ensures reproducibility, supports validity, and enables readers to assess susceptibility to bias slightly modified from STARD guidance. | Specifies statistical methods for accuracy (Se/Sp/PPV/NPV/LR/ROC), with model assumptions and CI computation: Diagnostic Accuracy Analysis The specificities and sensitivities of different SSEP change categories. SSEPs which had significant changes had a sensitivity of 30% (17%–44%) and a high specificity of 96% (94%–97%) (Table 3). Loss of waveforms had a sensitivity of 16% (5%–27%) and a specificity of 98% (97.5%–99%) (Table 3). The ROC curve for significant change of waveforms had an area under the curve (AUC) of 0.62, significance of 0.006, and 95% CI of 0.53 to 0.71. The AUC for loss of responses was 0.653 with an asymptotic significance of 0.019 and a 95% CI of 0.509 to 0.797. The AUC values for both significant changes and loss of responses to predict postoperative NDs are significant.  The odds ratio for significant changes in SSEPs was 9.80 with a 95% CI of 4.70 to 20.46. Loss of waveforms had an odds ratio of 11.82 with a 95% CI of 4.45 to 31.41. Using significant changes in SSEPs as the metric, there was 13/846 (1.54%) True Positives, 34/846 (4.02%) False Positives, 769/846 (90.9%) True Negatives, and 30/846 (3.5%) False Negatives. In terms of a loss of SSEPs, there was 7/846 (0.83%) True Positives, 11/846 (1.3%) False Positives, 792/846 (93.6%) True Negatives, and 39/846 (4.6%) False Negatives.(Thirumala et al., 2017)  Comparing IONM modalities: Based on this subgroup of studies, the absolute difference in the sensitivities of combined IONM and IONM with EEG alone was 20.0% (95% CI 6.1%–33.9%). The absolute difference in the false-positive rates of combined IONM and IONM with EEG alone was 7.0% (95% CI 5.6%–8.4%). The absolute difference in the sensitivities and specificities of combined IONM and IONM with SSEP alone were 10.0% (95% CI −0.8% to 20.8%) and 3.6% (95% CI 2.6%–4.6%), respectively. The McNemar test showed statistically significant evidence of differences in the sensitivities (Χ 2 (1df) = 6, P = 0.01) and false-positive rates (Χ 2 (1df) = 97, P = 0.001) of combined IONM and IONM with EEG alone. There was no statistically significant evidence of difference in the sensitivities (Χ 2 (1df) = 3, P = 0.08) of combined IONM and IONM with SSEP alone, whereas there was significant evidence of difference in the false-positive rates (Χ 2 (1df) = 50, P = 0.001) of these 2 techniques. For this subgroup, bimodal IONM, with a change in either EEG or SSEP as the designated alarm criterion, was 1.6 times more sensitive than exclusive use of EEG and 1.23 times as sensitive as independent SSEP monitoring.(Thiagarajan et al., 2015)  Alternate methos of IONM accuracy: Sensitivity, specificity, and their 95% confidence intervals are typically reported. Occasionally this reporting also includes likelihood ratios and subgroup analysis of IONM performance.(Holdefer et al., 2013) A hypothesis regarding acceptable performance is rarely reported. Receiver operating characteristic (ROC) curve analysis is frequently reported. Optimal thresholds derived from this analysis are often used in sensitivity and specificity calculations. (Muramoto et al., 2013) This data driven analysis (as compared to planned thresholds) can introduce bias. (Pan et al., 2024)  Methods for estimating IONM accuracy in the literature vary widely in their treatment of MEPs or SEPs that that recover with surgeon intervention. One method might be considered “context dependent” where an EP change in response to an adverse event that recovers with intervention is scored as a true positive. (Hilibrand et al., 2004) Another method is “end of surgery” that scores persistent EP changes to closing as positive test results and those that have recovered by closing negative test results.(Wilent et al., 2020) A third method simply omits recovered EPs from the analysis.(Kobayashi et al., 2014) A perhaps more conventional method for medical diagnosis considers any EP alert during surgery (recovered and persistent) as a positive test result.(Clark et al., 2013) A final approach bypasses traditional two by two contingency tables and consider recovered EP changes as a separate category. (Holdefer et al., 2016; Neuloh et al., 2004) Their association with outcomes is given but they are not included in conventional sensitivity and specificity calculations. In other studies, how recovered EP changes were scored is simply not reported.(Sutter et al., 2019) |
| 15 | How indeterminate index test or reference standard results were handled | In IONM studies, indeterminate results—such as borderline signal changes or technical failures—can significantly impact the assessment of diagnostic accuracy. To ensure transparency and reproducibility, it's essential to: Define and report the frequency and causes of indeterminate results. Specify handling methods, such as exclusion from analysis, categorization as a separate group, or reclassification as false positives/negatives based on clinical outcomes. Justify the chosen approach and discuss its potential impact on diagnostic accuracy estimates. Transparent reporting of these aspects aligns with the STARD 2015 guidelines, which emphasize the importance of addressing indeterminate results to avoid biased estimates of test performance. | Defines handling of indeterminate/partial-recovery cases (separate category vs collapsed) and reports sensitivity analyses: Indeterminate IONM results were divided into 2 main types of situations: 1) An IONM alert that normalized after corrective measures in a patient who emerged without new spinal cord deficits; it was difficult to determine whether such an event represented a true impending cord injury that was averted or a false-positive alert that spontaneously disappeared. 2) Delayed spinal cord injury; when the patient woke up after surgery, she demonstrated the same neurologic function as preoperatively but showed different levels of neurologic deficits after several hours of surgery.(Wang et al., 2018) |
| 16 | How missing data on the index test and reference standard were handled | Dealing with missing data is a challenging task. The most common approach is to exclude patients for whom the data of interest is missing. However, this approach is only valid if we have strong reasons to assume that the data is “missing at random” (MAR), meaning that the probability of missing is unrelated to unobserved data and does not introduce selection bias. For instance, if data is consistently missing for patients in the worst clinical condition because their condition prevents them from undergoing certain tests, excluding these patients would systematically omit the worst cases and introduce bias. Examples in IONM are in case of MEP loss, patients may not show up in the follow-up because they have a severe postoperative neurologic condition. Another issue is when the IONM team does not have access to follow-up patient charts, because of limited collaboration with the treating clinic. In such situations, it is highly recommended to consult a statistician to determine the most appropriate method for handling missing data. Alternatives to simple exclusion exist, such as data imputation (e.g. complete with the population mean, assign an existing value, etc.), which can be used to retain information from patients with incomplete data and reduce potential bias. The methods used to deal with missing data should be stated explicitly in the text, or at the very least in the limitations of the study. | Describes missing-data mechanisms and handling (complete-case vs imputation), mitigating bias: In the study "Evaluation of intraoperative neuromonitoring (IONM) data with the Mainz IONM Quality Assurance and Analysis tool," the handling of missing data was meticulously documented. Starting with 1935 IONM data files, 1921 were initially readable, with 14 files excluded due to unreadable formats. From these, 34 files were further excluded for missing data labelling, ensuring systematic identification of incomplete records. During the initial cleaning phase, 56 files were flagged for exclusion, but a manual review recovered 42 of them, resulting in 1879 files for analysis. Automated plausibility checks then identified potential labelling errors or inconsistencies in 1138 files, representing 60% of the analyzed dataset. Of these, 915 files, or 48.5%, were confirmed erroneous and manually corrected, involving specific actions such as adding 1012 labels, modifying 916 labels, reordering 102 labels, and deleting 868 labels. These corrections totaled 2898 changes, accounting for 16% of the labelled EMG data. The process was supported by the Mainz IONM Quality Assurance tool, accessible at ionmreference.net, which facilitated data review and editing, ensuring a robust and transparent approach to managing missing and erroneous data.(Musholt et al., 2023) |
| 17 | Any analyses of variability in diagnostic accuracy, distinguishing pre-specified from exploratory | If the variance in the entire population is high, it might be a good idea to consider subgroup analyses. Possible explanations for high variances could be Simpson’s paradox (when subgroups have different trends than the entire population) or the presence of outliers (beware also of typos!). Outliers can for instance be detected via box plots with whiskers. A stratification should be considered and justified. For example, if patients have preoperative neurologic deficits or motor weaknesses, this might influence the MEPs and the distribution of the outcome data. It might therefore be reasonable to stratify the results according to this criterion.  It is crucial to distinguish pre-specified analyses from exploratory analyses. Before any data collection, a main hypothesis should be formulated, defining a primary endpoint (or at most two, but ideally not more). Based on this hypothesis, an appropriate statistical test should be selected to address the question raised. This approach enables a power analysis (i.e., determining the required sample size) and ensures that the full statistical power is dedicated to this test, increasing the likelihood of detecting a significant result if one exists. | Predefines variability/subgroup analyses (e.g., modality, procedure, anaesthesia) and distinguishes exploratory work: The study focused on IONM during thyroid surgery, with the index test being IONM signals (e.g., electromyogram for RLN function) and the reference standard being postoperative RLN injury assessed by laryngoscopy. It included 500 patients (1000 RLNs at risk) using NIM 3.0 Response equipment. Pre-specified Analyses and Standardization: The researchers adhered to a standardized protocol recommended by the International Neural Monitoring Study Group (INMSG), employing a predefined criterion for loss of signal (LOS) as a V2 amplitude ≤100 μV. This pre-specified threshold facilitated the calculation of diagnostic accuracy metrics, yielding a sensitivity of 92.0%, specificity of 99.3%, positive predictive value (PPV) of 76.7%, and negative predictive value (NPV) of 99.8%. Exploratory Analyses and ROC Curve Evaluation: Beyond the pre-specified analyses, the study conducted exploratory evaluations using receiver operating characteristic (ROC) curve analysis to identify an optimal cutoff value for predicting postoperative vocal fold paresis. This analysis suggested that a V2 amplitude ≤189 μV might offer improved prognostic value, with a PPV of 77.4% and an NPV of 99.9%. The authors clearly distinguished this exploratory finding from the pre-specified analyses, emphasizing the need for cautious interpretation and further validation in multicenter studies. Consideration of Variability and Stratification: "Publications emphasize high variability in predictive values, with positive result (loss of signal, LOS) ranging from 10–90% and negative result (preservation of normal signal) ranging from 70–99%." Furthermore, the paper compares its results with specific studies, such as Genther et al., which reported PPV 72.4% and NPV 99.9% for V2 amplitude <200 μV, as part of this discussion. The text mentions: "Comparative studies, such as Genther et al. (V2 amplitude <200 μV): PPV 72.4%, NPV 99.9% [18]," further illustrating variability.(Stopa and Barczyński, 2017) |
| 18 | Intended sample size and how it was determined | Sample size calculations are crucial in diagnostic accuracy studies to ensure sufficient precision and align with study objectives and hypotheses. Report when and how the sample size was determined and whether the assumptions made were in line with the study objective | Reports sample-size rationale linked to precision of accuracy estimates: “The sample size was estimated based on the principle of detecting a 5 % difference in the incidence of primary or secondary outcome measures with a 90 % probability at p < 0.05. The incidence of nerve events was calculated based on the number of nerves at risk. The analysis of both primary and secondary outcomes was performed on an intention-to-treat basis(Barczyński et al., 2012). “ |
| RESULTS | | | |
| Participants | | | |
| 19 | Report flow of participants, using a diagram. | A participant flow diagram provides a visual summary of the progression of individuals through the stages of a study. It typically includes the number of participants assessed for eligibility, excluded (with reasons), enrolled, allocated to interventions, followed up, and analyzed. In the context of IONM, such diagrams can illustrate the number of patients undergoing monitoring, those with successful signal acquisition, instances of signal loss, and correlations with postoperative outcomes. | Provides a flow diagram with numbers at each stage, clarifying attrition and verification: ”Patient IONM characteristics are reported in Fig. 1. Among patients with TIA, 9 patients (19.6%) displayed significant IONM changes during their CEA. Among these patients with TIA, 7 showed significant dual modality changes, consisting of 7 permanent SSEP changes, 4 transient EEG changes, and 3 permanent EEG changes. One patient displayed only a transient SSEP change, and one patient displayed only a transient EEG change. In total, 37 patients (80.4%) with TIA did not display IONM changes. Among 2186 patients without TIA, 236 patients (10.8%) displayed significant IONM changes during their CEA. Among these patients without TIA, 95 showed significant SSEP (160 transient and 11 permanent) and EEG (142 transient and 18 permanent) changes, 76 patients displayed only SSEP changes (69 transient and 7 permanent), and 65 patients displayed only EEG changes (56 transient and 9 permanent). In total, 1950 patients (89.2%) without TIA did not display IONM changes, though 55 (2.5%) of these patients were recorded as having a stroke.(Moehl et al., 2022)” |
| 20 | Baseline demographic and clinical characteristics of participants | Report baseline demographic and clinical characteristics relevant to IONM interpretation, including age, sex, diagnosis, preoperative neurologic status, and relevant surgical features  In intraoperative neurophysiological monitoring (IONM) studies, reporting baseline demographic and clinical characteristics—such as age, sex, and preoperative neurological status—is critical. These factors can influence evoked potential amplitudes, signal reliability, and the likelihood of intraoperative alerts or false positives. For example, older patients or those with preexisting motor deficits may have lower baseline amplitudes or delayed responses, impacting both the index test and outcome classification. Detailed characterization of the surgical population also helps assess whether findings are generalizable to other settings and whether diagnostic accuracy measures (sensitivity, specificity, NPV, PPV) apply equally across patient subgroups. This information supports valid interpretation of IONM performance and helps identify sources of variability. Report anesthesia context relevant to interpretation: technique, depth monitor values at first acquisition, NMB level before MEPs, mean MAP/EtCO₂/core temperature during IONM. | Presents baseline demographics/clinical features relevant to IONM signal interpretation: “Results: We included 100 patients; their mean age was 48.47 ± 13.81 years, and 61 % were female. Of the 100 participants, 50 were diagnosed with vestibular schwannoma, 20 with posterior fossa meningioma, 11 with clivus chordoma, and 19 with other posterior fossa pathologies (see Table 1). The median HB grade prior to surgery was I (range I-IV). Ninety-three patients had normal facial function prior to surgery (HB I), six had slight/moderate facial paresis (HB II/III), and one had a moderate-severe facial paresis (patient #30) (HB IV) Behavior of Facial corticobulbar motor evoked potentials: The monitorability rate of patients with normal facial nerve function (HB I) or slight/moderate (HB II/III) facial paresis was 100 %. In all patients but one with moderately severe facial nerve paralysis (HB IV), we elicited FCoMEP regardless of the preoperative facial nerve status. The patient in whom we could not elicit responses presented moderately severe facial paresis (HB IV) before surgery and ended up with total facial paralysis (HB VI). One patient presented moderate facial paresis and no significant decrease in amplitude of FCoMEP at the end of surgery, so this was considered a false negative.(Fernández-Conejero et al., 2022)” |
| 21 | Distribution of severity of disease in those with the target condition | Report the distribution of severity of the target condition, including degree, duration, and permanence of postoperative neurologic deficit where applicable.  In studies using intraoperative neurophysiological monitoring (IONM) to detect or predict postoperative neurological deficits, the target condition can vary in severity—from mild, transient changes to severe, permanent impairments. Reporting this distribution is essential because the sensitivity of IONM may be higher in more severe cases, where signal changes are more pronounced. Stratifying outcomes by degree and duration of deficit allows a more accurate assessment of how IONM performs across the clinical spectrum.  Equally important is the characterization of postoperative findings in patients who do not meet the definition of a neurological deficit. These individuals may still present with transient symptoms, subjective complaints (e.g., numbness, paresthesia), or unrelated complications that may account for intraoperative IONM changes. Describing the range and frequency of these alternative outcomes provides crucial context for interpreting false positives and evaluating the test’s specificity. Without this information, test performance may be misestimated. Together, these data allow for a realistic, nuanced understanding of IONM accuracy and generalizability in surgical practice. | Reports disease severity distribution in target-condition cases, informing spectrum bias: “In this study of 238 patients undergoing microvascular decompression (MVD) with intraoperative BSER monitoring, postoperative hearing outcomes were analyzed based on intraoperative changes in wave V. Among the 35 patients who experienced a complete loss of wave V, 15 had persistent hearing loss, with 9 (3.8%) exhibiting poor hearing (grades C/D) and 6 (2.5%) retaining good hearing (grades A/B). The remaining 20 had transient loss, with 5 (2.1%) showing hearing grade C/D and 15 (6.3%) grade A/B. Thus, 14 of 35 patients (40%) in this group had poor hearing outcomes. In the group with significant changes in wave V (n = 49), postoperative audiograms revealed that 5 patients (2.1%) had hearing grade C/D while 44 (18.4%) retained hearing grade A/B. Among the 154 patients who showed no change in wave V, only 2 (0.8%) had hearing grade C/D and the vast majority, 152 (63.8%), maintained good hearing. These results demonstrate a clear correlation between intraoperative wave V changes and postoperative hearing, with the highest risk of hearing loss observed in those with complete wave V loss and the lowest in those with no change.(P.D. Thirumala et al., 2014)” |
| 22 | Time interval and any clinical interventions between index test and reference standard | Report the time interval between the index test and the reference standard, and describe any clinical interventions performed in response to IONM changes. Specify the timing of baseline acquisition, the last IONM recording relative to closure, and the timing of anesthetic, physiologic, or surgical interventions after alerts  In IONM studies, the index test—typically a neurophysiological alert during surgery—is often compared to a reference standard such as a postoperative neurological exam or longer-term follow-up. It is essential to report the time interval between the index test and the outcome assessment, as changes in the patient’s condition may occur due to natural recovery, delayed injury, or interventions performed after the alert. These changes can bias the relationship between the index test and the reference standard, particularly if IONM alerts lead to immediate surgical or anesthetic maneuvers that prevent a deficit from manifesting. Therefore, studies should describe not only the timing of follow-up (e.g., immediate post-op, discharge, or 3-month visit), but also any interventions performed in response to IONM alerts, such as stopping dissection, increasing blood pressure, or administering steroids. These actions may influence the outcome and can obscure whether the index test truly predicted the deficit or contributed to its avoidance. Some have proposed using the immediate intraoperative resolution of an IONM alert as an alternative or secondary reference standard, which may provide a closer temporal link and reduce post-surgical bias. Regardless of the model used, transparent reporting of both the follow-up interval and all relevant clinical interventions is critical to interpret the true diagnostic value of IONM. Report time of baseline and time of last IONM recording relative to skin closure, plus timestamps for anesthetic/physiologic interventions after alerts. | Reports time of baseline and last intra-op acquisition per modality, plus alert→intervention→final-status timings—reducing risk of late false negatives.: "In the other 5 patients harboring a tumor involving the dorsal portion of the premotor cortex and the upper insula, LFBS (2.5–5 mA) induced ADs in 2 patients on stimulating the ventral premotor cortex; therefore, HFMS (8–10 mA, 1 Hz) was used. The HFMS effectively induced anarthria without seizure activity. No paraphasias or anomias were observed in these 5 patients during the naming task when HFMS with a repetition rate of 1 Hz was applied to other sites of the frontal and temporal cortex. An increase in the train repetition rate from 1 to 3 Hz and the same current intensity (8–10 mA) led to the identification of cortical sites determining paraphasias and anomias. Electromyography detected no evoked muscle potential during these language disturbances. This adjustment of the stimulation frequency enabled the identification of essential cortical language sites over the frontal cortex as well as the surgical point of entry. When applied with a repetition rate of 3 Hz at the subcortical level, the HFMS induced phonemic and semantic paraphasias and thus identified as functional boundaries, respectively, the dorsal language tracts (that is, the arcuate and superior longitudinal fascicles) and the inferior frontooccipital fascicle, with no MEPs in these sites." (Riva et al., 2016)  "IONM Changes, Surgical Strategy, and Clinical Correlations: Overall significant IONM changes (at least 1 evoked potential modality) were registered in 14 (12.96%) of the 108 patients. In 5 patients (4.62%), these events resolved to baseline levels after temporary stop of surgery and gross-total resection could be achieved (stop-and-go surgery). In the other 9 patients (8.33%), we noted permanent loss of at least one of the 3 evoked potentials. However, only in 2 cases (1.85%) were the SSEPs and MEPs lost, and the D-wave permanently dropped by about 50%. In these 2 cases, after several attempts to restart the surgery with a change in strategy, the operation was definitively abandoned to prevent permanent paraplegia (stop surgery), and a residual tumor was left in place. These patients, described in our previous study on IONM,7 presented with a persistent new motor deficit at the 6-month follow-up. However, these patients recovered their preoperative neurological status at the 12-month follow-up. In the other 7 patients (7.40%), the D-wave was stable or decreased no more than 50% in combination with alteration of MEPs and/or SSEPs and allowed us to the continue surgery, because neurophysiological literature suggests that patients usually wake up after the operation with a motor deficit but invariably recover over a period of days or weeks. Among these patients, 4 emerged from surgery with a new transitory motor deficit while the other 3 patients had no postoperative deficit (false positive)."(Ghadirpour et al., 2019) |
| Test results | | | |
| 23 | Cross tabulation of the index test results (or their distribution) by the results of the reference standard | A clear cross-tabulation, typically in a 2×2 table—or a prespecified table should be provided, displaying the number of true positives, false positives, false negatives, and true negatives, based on the index test (e.g., IONM signal changes) and the reference standard (e.g., postoperative neurological outcome). This format allows calculation of sensitivity, specificity, PPV, and NPV, and supports transparent interpretation of the test's diagnostic accuracy.  IONM-specific considerations:  In IONM studies, the index test may be defined as a significant change in evoked potentials (e.g., >50% amplitude drop), while the reference standard is often a postoperative neurological deficit. Given the clinical implications of real-time intraoperative decisions, providing a complete cross-tabulation enables readers to assess both performance and potential biases in outcome classification. Extended tables may also be used when multiple thresholds or outcomes are assessed.  If reversible changes are separated, consider an expanded 3×2 (ND/RD/ID × deficit/no-deficit); state how RD/partial recovery was classified (if available) and show sensitivity analyses for its impact on Se/Sp/PPV/NPV/AUC. | Provides clear cross-tabs (per structure or per patient) linking index-test categories to the reference outcome.: "In a study involving 326 cases, the diagnostic accuracy of various intraoperative neurophysiological monitoring (IONM) changes was evaluated using 2×2 tables. For SSEP changes alone, there were 20 true positives (TP), 24 false positives (FP), 275 true negatives (TN), and 7 false negatives (FN). MEP changes yielded 24 TP, 33 FP, 266 TN, and 3 FN. When either SSEP or MEP changes were considered (i.e., a positive result if either modality showed a change), the analysis showed 26 TP, 42 FP, 257 TN, and only 1 FN. Conversely, requiring both SSEP and MEP changes to be present resulted in 18 TP, 15 FP, 284 TN, and 9 FN. (Liu et al., 2022) "lower extremity (LE) significant changes, there was 1/1057 (0.09%) True Positives, 8/1057 (0.75%) False Positives, 1013/1057 (95.8%) True Negatives, and 35/1057 (3.3%) False Negatives. Per LE loss of waveforms, there were 1/1057 (0.09%) True Positives, 1/1057 (0.09%) False Positives, 1020/1057 (96.5%) True Negatives, and 35/1057 (3.3%) False Negatives."(Hiruta et al., 2021) |
| 24 | Estimates of diagnostic accuracy and their precision | Report estimates of diagnostic accuracy with measures of precision (for example, 95% confidence intervals), and state how RD and PR were handled in the calculations. If sensitivity analyses were performed to show the impact of alternate classifications, report them.  Diagnostic accuracy studies should present quantitative estimates—such as sensitivity, specificity, positive predictive value (PPV), negative predictive value (NPV), and odds ratios—along with measures of statistical precision, including 95% confidence intervals. P-values may be included where appropriate, though effect size and precision are more informative. These statistics enable critical appraisal of the reliability and clinical significance of the IONM modality under study. IONM-specific considerations: In IONM research, where sample sizes may be limited or outcomes infrequent, it's essential to report these measures transparently. If statistical estimates are not provided, authors should clearly explain why (e.g., exploratory study, limited sample). Precision measures are particularly important given the variability in intraoperative responses and outcome definitions. State how reversible deterioration/partial recovery was handled in accuracy calculations (separate category vs collapsed to ±) and provide sensitivity analyses showing its impact on Se/Sp/PPV/NPV and AUC. Report 95% CIs and model/test used for all estimates. | Reports accuracy estimates with 95% CIs and, if applicable, category-expanded summaries (e.g., PR/RD/ID): The diagnostic performance of various modalities for detecting changes in evoked potentials (EP), including motor evoked potentials (MEP) and somatosensory evoked potentials (SSEP), was evaluated in a comprehensive analysis. The modality assessing changes in any EP (MEP or SSEP) demonstrated a sensitivity of 90.0% (95% CI: 59.6%–99.5%) and specificity of 93.9% (95% CI: 90.6%–96.0%), with a positive predictive value (PPV) of 32.1% (95% CI: 17.9%–50.7%) and negative predictive value (NPV) of 99.7% (95% CI: 98.1%–100.0%). (Park et al., 2021) |
| 25 | Any adverse events from performing the index test or the reference standard | Although IONM techniques are generally considered low risk, adverse events may occur due to electrical stimulation, electrode placement, or equipment malfunction. In IONM, index test-related events include tongue lacerations, dental trauma, thermal burns from electrodes, and inadvertent patient movement during stimulation. Reference standard-related events may involve additional imaging, neurological testing, or interventions postoperatively. Accurate reporting of such adverse events is essential to assess the safety profile of IONM and refine monitoring protocols. | Explicitly lists adverse events attributable to index or reference tests, with frequency and severity: No complications (e.g., bite injuries of the tongue and the inner cheeks or infections at the insertion site of the corkscrew or subdermal needle electrodes) were seen. In one patient, a single intraoperative seizure occurred at the beginning of the surgery while the mMEP baseline recordings elicited by TES were being taken. In this patient, further anesthesia management included pentobarbital administration. This patient's medical history was remarkable for symptoms resembling an SAH 2 years before surgery without definite diagnosis and a human immunodeficiency virus infection. There was no history of seizure.(Szelényi et al., 2006) |
| DISCUSSION | | | |
| 26 | Study limitations, including sources of potential bias, statistical uncertainty, and generalizability | Definition of Test Criteria Clearly describe the neurophysiological test criteria (e.g., specific warning criteria used, cut-off thresholds for alarms). Specify timing of outcome assessment (e.g., immediate post-op, days after surgery, or long-term follow-up). Statistical Methods For small samples, justify statistical test selection based on assumptions (e.g., chi-square requires ≥5 expected per cell). For very small samples (<12), avoid inferential statistics; instead use descriptive statistics and acknowledge limitations. Correction for Multiple Testing Indicate whether multiple testing correction was performed. Specify method used (e.g., Bonferroni, Holm–Bonferroni). If no correction applied, justify and discuss implications. Adjustment for Confounders Report if multivariable regression or other adjustment methods were used to control for confounders. If not adjusted, acknowledge and include in limitations section.  Discuss limitations from anesthetic variability (technique shifts, depth drift, NMB fluctuations) and unrecorded physiologic changes as potential accuracy bias. | Transparently discusses bias sources (selection, verification, incorporation), uncertainty, and generalizability. “This prospective multicenter cohort study enrolled 1,934 patients across 16 Japanese spine centers to validate the alarm point for IONM using transcranial electrical stimulation muscle motor evoked potentials (Tc(E)-MEPs).(Takahashi et al., 2021) Alarm Threshold Definition: The study explicitly defined the alarm point as a ≥70% decrease in Tc(E)-MEP amplitude. This threshold was formulated by the Monitoring Working Group of the Japanese Society for Spine Surgery and Related Research (JSSR) and aimed to standardize IONM alerts across various spinal surgeries. Outcome Assessment Timing: Postoperative neurological deficits were assessed to determine the efficacy of the alarm threshold. The study categorized patients into high-risk (Group HR) and common surgery (Group C) groups, allowing for stratified analysis based on surgical risk factors. Statistical Analysis and Sample Size Justification: The large sample size enabled robust statistical analysis. In Group HR, the sensitivity and specificity were 94.4% and 87.0%, respectively. In contrast, Group C exhibited a sensitivity of 63.6% and specificity of 91.9%. The statistically significant lower sensitivity in Group C (P < 0.05) highlighted the need for cautious application of the alarm threshold in lower-risk surgeries. Adjustment for Confounders and Limitations: The study acknowledged that the alarm point's validity might vary across different surgical contexts. It emphasized the necessity for careful interpretation of IONM alerts, especially in surgeries not classified as high-risk. By stratifying patients and analyzing subgroups, the study addressed potential confounding factors and provided a nuanced understanding of the alarm threshold's applicability. |
| 27 | Implications for practice, including the intended use and clinical role of the index test | The clinical relevance of intraoperative neurophysiological monitoring (IONM) depends on differences in baseline characteristics—such as clinical status and neurophysiological data—between study populations and the patients typically encountered in surgical settings.  Key considerations should include: (1) the stage of development of IONM, ranging from proof-of-concept validation to accuracy assessment in specific surgical populations, evaluation in the broader target population, and demonstration of its incremental benefit in routine clinical use; and (2) the additional research required to optimize its application for real-world surgical practice. Emphasizing these aspects clarifies IONM’s clinical utility, its role in intraoperative decision-making, and its potential to enhance patient safety and surgical outcomes. | Interprets accuracy in clinical context, specifies intended use, and outlines implications/limits for practice: ‘These preliminary results show that it is feasible to record the Blink reflex in patients under general anesthesia using low doses of desflurane or sevoflurane during surgery if a short train of electrical stimuli is applied to the supraorbital nerve.(Deletis et al., 2009) 2.Bulbocavernous Reflex (BCR) had low sensitivity for voiding dysfunction at discharge but had high accuracy at 6-month follow-up examinations. BCR loss was associated with new voiding dysfunction. Intraoperative BCR monitoring is a potentially useful tool for enhancing safety during posterior lumbar fusion by predicting postoperative voiding dysfunction.(Choi et al., 2022)  3. In children whose spinal deformity surgery was aborted due to intraoperative IONM loss, there was a strong correlation between combined intraoperative SSEP/MEP loss, the magnitude of IONM loss, the timing of clinical recovery, and the time of electrophysiological IONM recovery. The highest likelihood of having a prolonged postoperative neurological deficit and undetectable IONM signals upon return to the OR occurs with bilateral complete loss of SSEPs and MEPs.(CreveCoeur et al., 2024)” |
| OTHER INFORMATION | | | |
| 28 | Registration number and name of registry | Provide trial/registry name and registration number (or state “not registered”) and give the registration URL if available. |  |
| 29 | Where the full study protocol can be accessed | Provide a public link or repository identifier for the full protocol (or state unavailability) and summarize deviations from protocol. |  |
| 30 | Sources of funding and other support; role of funders | Disclose funding sources and other support; describe the role of funders/sponsors in design, conduct, analysis, and reporting. |  |

Read with Table 2 (STARD-IONM checklist) and Supplementary Table 2 (Anesthesia elements)

References:

Al-Qudah AM, Sivaguru S, Anetakis K, Crammond DJ, Balzer JR, Thirumala PD, et al. Role of Intraoperative Electroencephalography in Predicting Postoperative Delirium in Patients Undergoing Cardiovascular Surgeries. Clin Neurophysiol 2024;164:40–6. https://doi.org/10.1016/j.clinph.2024.05.012.

Barczyński M, Konturek A, Stopa M, Honowska A, Nowak W. Randomized Controlled Trial of Visualization versus Neuromonitoring of the External Branch of the Superior Laryngeal Nerve during Thyroidectomy. World J Surg 2012;36:1340–7. https://doi.org/10.1007/s00268-012-1547-7.

Boaro A, Azzari A, Basaldella F, Nunes S, Feletti A, Bicego M, et al. Machine learning allows expert level classification of intraoperative motor evoked potentials during neurosurgical procedures. Comput Biol Med 2024;180:109032. https://doi.org/10.1016/j.compbiomed.2024.109032.

Calancie B, Donohue ML, Moquin RR. Neuromonitoring with pulse-train stimulation for implantation of thoracic pedicle screws: a blinded and randomized clinical study. Part 2. The role of feedback. J Neurosurg Spine 2014;20:692–704. https://doi.org/10.3171/2014.2.SPINE13649.

Choi J, Kim J-S, Hyun S-J, Kim K-J, Kim H-J, Deletis V, et al. Intraoperative bulbocavernosus reflex monitoring in posterior lumbar fusion surgery. Clin Neurophysiol 2022;144:59–66. https://doi.org/10.1016/j.clinph.2022.09.020.

Clark AJ, Ziewacz JE, Safaee M, Lau D, Lyon R, Chou D, et al. Intraoperative neuromonitoring with MEPs and prediction of postoperative neurological deficits in patients undergoing surgery for cervical and cervicothoracic myelopathy. Neurosurg Focus 2013;35:E7. https://doi.org/10.3171/2013.4.FOCUS13121.

CreveCoeur TS, Iyer RR, Goldstein HE, Delgardo MW, Hankinson TC, Erickson MA, et al. Timing of intraoperative neurophysiological monitoring (IONM) recovery and clinical recovery after termination of pediatric spinal deformity surgery due to loss of IONM signals. Spine J 2024;24:1740–9. https://doi.org/10.1016/j.spinee.2024.04.008.

Deletis V, Urriza J, Ulkatan S, Fernandez‐Conejero I, Lesser J, Misita D, et al. The feasibility of recording blink reflexes under general anesthesia. Muscle Nerve 2009;39:642–6. https://doi.org/10.1002/mus.21257.

Dulfer SE, Gadella MC, Tamási K, Absalom AR, Lange F, Scholtens-Henzen CHM, et al. Use of NEedle Versus suRFACE Recording Electrodes for Detection of Intraoperative Motor Warnings: A Non-Inferiority Trial. The NERFACE Study Part II. J Clin Med 2023;12:1753. https://doi.org/10.3390/jcm12051753.

Dulfer SE, Sahinovic MM, Lange F, Wapstra FH, Postmus D, Potgieser ARE, et al. The influence of depth of anesthesia and blood pressure on muscle recorded motor evoked potentials in spinal surgery. A prospective observational study protocol. J Clin Monit Comput 2021;35:967–77. https://doi.org/10.1007/s10877-020-00645-1.

Fernández-Conejero I, Ulkatan S, Sen C, Miró Lladó J, Deletis V. Intraoperative monitoring of facial corticobulbar motor evoked potentials: Methodological improvement and analysis of 100 patients. Clin Neurophysiol 2022;142:228–35. https://doi.org/10.1016/j.clinph.2022.08.006.

Ghadirpour R, Nasi D, Iaccarino C, Romano A, Motti L, Sabadini R, et al. Intraoperative neurophysiological monitoring for intradural extramedullary spinal tumors: predictive value and relevance of D-wave amplitude on surgical outcome during a 10-year experience. J Neurosurg Spine 2019;30:259–67. https://doi.org/10.3171/2018.7.SPINE18278.

Greve T, Wang L, Katzendobler S, Geyer LL, Schichor C, Tonn JC, et al. Bilateral and Optimistic Warning Paradigms Improve the Predictive Power of Intraoperative Facial Motor Evoked Potentials during Vestibular Schwannoma Surgery. Cancers (Basel) 2021;13:6196. https://doi.org/10.3390/cancers13246196.

Hilibrand AS, Schwartz DM, Sethuraman V, Vaccaro AR, Albert TJ. Comparison of transcranial electric motor and somatosensory evoked potential monitoring during cervical spine surgery. J Bone Jt Surg - Ser A 2004. https://doi.org/10.2106/00004623-200406000-00018.

Hiruta R, Sato T, Itakura T, Fujii M, Sakuma J, Bakhit M, et al. Intraoperative transcranial facial motor evoked potential monitoring in surgery of cerebellopontine angle tumors predicts early and late postoperative facial nerve function. Clin Neurophysiol 2021;132:864–71. https://doi.org/10.1016/j.clinph.2020.12.025.

Holdefer RN, Heffez DS, Cohen BA. Utility of Evoked EMG Monitoring to Improve Bone Screw Placements in the Cervical Spine. J Spinal Disord Tech 2013;26:E163–9. https://doi.org/10.1097/BSD.0b013e31828871a1.

Holdefer RN, MacDonald DB, Guo L, Skinner SA. An evaluation of motor evoked potential surrogate endpoints during intracranial vascular procedures. Clin Neurophysiol 2016;127:1717–25. https://doi.org/10.1016/j.clinph.2015.09.133.

Khan MH, Smith PN, Balzer JR, Crammond D, Welch WC, Gerszten P, et al. Intraoperative somatosensory evoked potential monitoring during cervical spine corpectomy surgery: Experience with 508 cases. Spine (Phila Pa 1976) 2006. https://doi.org/10.1097/01.brs.0000200163.71909.1f.

Kim MH, Park J, Park YG, Cho YE, Kim D, Lee DJ, et al. Comparison of intraoperative neurophysiological monitoring between propofol and remimazolam during total intravenous anesthesia in the cervical spine surgery: a prospective, double-blind, randomized controlled trial. Korean J Anesthesiol 2025;78:16–29. https://doi.org/10.4097/kja.24613.

Kobayashi S, Matsuyama Y, Shinomiya K, Kawabata S, Ando M, Kanchiku T, et al. A new alarm point of transcranial electrical stimulation motor evoked potentials for intraoperative spinal cord monitoring: a prospective multicenter study from the Spinal Cord Monitoring Working Group of the Japanese Society for Spine Surgery and Related. J Neurosurg Spine 2014;20:102–7. https://doi.org/10.3171/2013.10.SPINE12944.

Korn A, Halevi D, Lidar Z, Biron T, Ekstein P, Constantini S. Intraoperative neurophysiological monitoring during resection of intradural extramedullary spinal cord tumors: experience with 100 cases. Acta Neurochir (Wien) 2015;157:819–30. https://doi.org/10.1007/s00701-014-2307-2.

Kundnani VK, Zhu L, Tak HH, Wong HK. Multimodal intraoperative neuromonitoring in corrective surgery for adolescent idiopathic scoliosis: Evaluation of 354 consecutive cases. Indian J Orthop 2010;44:64–72. https://doi.org/10.4103/0019-5413.58608.

Liu T, Yan L, Qi H, Luo Z, Liu X, Yuan T, et al. Diagnostic Value of Multimodal Intraoperative Neuromonitoring by Combining Somatosensory-With Motor-Evoked Potential in Posterior Decompression Surgery for Thoracic Spinal Stenosis. Front Neurosci 2022;16. https://doi.org/10.3389/fnins.2022.879435.

Malcharek MJ, Kulpok A, Deletis V, Ulkatan S, Sablotzki A, Hennig G, et al. Intraoperative Multimodal Evoked Potential Monitoring During Carotid Endarterectomy. Anesth Analg 2015;120:1352–60. https://doi.org/10.1213/ANE.0000000000000337.

McDevitt WM, Quinn L, Wimalachandra WSB, Carver E, Stendall C, Solanki GA, et al. Amplitude-reduction alert criteria and intervention during complex paediatric cervical spine surgery. Clin Neurophysiol Pract 2022;7:239–44. https://doi.org/10.1016/j.cnp.2022.07.003.

Moehl K, Shandal V, Anetakis K, Paras S, Mina A, Crammond D, et al. Predicting transient ischemic attack after carotid endarterectomy: The role of intraoperative neurophysiological monitoring. Clin Neurophysiol 2022;141:1–8. https://doi.org/10.1016/j.clinph.2022.06.010.

Muramoto A, Imagama S, Ito Z, Wakao N, Ando K, Tauchi R, et al. The cutoff amplitude of transcranial motor-evoked potentials for predicting postoperative motor deficits in thoracic spine surgery. Spine (Phila Pa 1976) 2013;38:E21-7. https://doi.org/10.1097/BRS.0b013e3182796b15.

Musholt TJ, Staubitz JI, Musholt PB. Evaluation of intraoperative neuromonitoring (IONM) data with the Mainz IONM Quality Assurance and Analysis tool. BJS Open 2023;7. https://doi.org/10.1093/bjsopen/zrad051.

Neuloh G, Pechstein U, Cedzich C, Schramm J. Motor evoked potential monitoring with supratentorial surgery. Neurosurgery 2004;54:1061–70; discussion 1070-2. https://doi.org/10.1227/01.neu.0000119326.15032.00.

Ohy JB, Formentin C, Gripp DA, Nicácio Jr JA, Velho MC, Vilany LN, et al. Filling the gap: brief neuropsychological assessment protocol for glioma patients undergoing awake surgeries. Front Psychol 2024;15. https://doi.org/10.3389/fpsyg.2024.1417947.

Pan S-Y, Holdefer RN, Wu H-L, Li C-R, Guo L. The predictive value of intraoperative facial motor evoked potentials in cerebellopontine angle tumor surgery. Clin Neurophysiol 2024;166:176–90. https://doi.org/10.1016/j.clinph.2024.07.021.

Park D, Kim BH, Lee S-E, Jeong E, Cho K, Park JK, et al. Usefulness of Intraoperative Neurophysiological Monitoring During the Clipping of Unruptured Intracranial Aneurysm: Diagnostic Efficacy and Detailed Protocol. Front Surg 2021;8. https://doi.org/10.3389/fsurg.2021.631053.

Raabe A, Beck J, Schucht P, Seidel K. Continuous dynamic mapping of the corticospinal tract during surgery of motor eloquent brain tumors: evaluation of a new method. J Neurosurg 2014;120:1015–24. https://doi.org/10.3171/2014.1.JNS13909.

Rho YJ, Rhim SC, Kang JK. Is intraoperative neurophysiological monitoring valuable predicting postoperative neurological recovery? Spinal Cord 2016;54:1121–6. https://doi.org/10.1038/sc.2016.65.

Riva M, Fava E, Gallucci M, Comi A, Casarotti A, Alfiero T, et al. Monopolar high-frequency language mapping: can it help in the surgical management of gliomas? A comparative clinical study. J Neurosurg 2016;124:1479–89. https://doi.org/10.3171/2015.4.JNS14333.

Schwartz SL, Kale EB, Madden D, Husain AM. “Quadripolar” Transcranial Electrical Stimulation for Motor Evoked Potentials. J Clin Neurophysiol 2022;39:92–7. https://doi.org/10.1097/WNP.0000000000000751.

Seidel K, Beck J, Stieglitz L, Schucht P, Raabe A. The warning-sign hierarchy between quantitative subcortical motor mapping and continuous motor evoked potential monitoring during resection of supratentorial brain tumors. J Neurosurg 2013;118:287–96. https://doi.org/10.3171/2012.10.JNS12895.

Seidel K, Wermelinger J, Alvarez-Abut P, Deletis V, Raabe A, Zhang D, et al. Cortico-cortical evoked potentials of language tracts in minimally invasive glioma surgery guided by Penfield stimulation. Clin Neurophysiol 2024;161:256–67. https://doi.org/10.1016/j.clinph.2023.12.136.

Siller S, Szelényi A, Herlitz L, Tonn JC, Zausinger S. Spinal cord hemangioblastomas: significance of intraoperative neurophysiological monitoring for resection and long-term outcome. J Neurosurg Spine 2017;26:483–93. https://doi.org/10.3171/2016.8.SPINE16595.

Silverstein JW, D’Amico RS, Mehta SH, Gluski J, Ber R, Sciubba DM, et al. The diagnostic accuracy of neuromonitoring for detecting postoperative bowel and bladder dysfunction in spinal oncology surgery: a case series. J Neurooncol 2024;169:409–22. https://doi.org/10.1007/s11060-024-04742-y.

Squintani G, Basaldella F, Badari A, Rasera A, Tramontano V, Pinna G, et al. Intraoperative Neurophysiological Monitoring in Tethered Cord Syndrome Surgery: Predictive Values and Clinical Outcome. J Clin Neurophysiol 2025;42:257–63. https://doi.org/10.1097/WNP.0000000000001096.

Stopa M, Barczyński M. Prognostic value of intraoperative neural monitoring of the recurrent laryngeal nerve in thyroid surgery. Langenbeck’s Arch Surg 2017;402:957–64. https://doi.org/10.1007/s00423-016-1441-0.

Sultan I, Bianco V, Kilic A, Jovin T, Jadhav A, Jankowitz B, et al. Predictors and Outcomes of Ischemic Stroke After Cardiac Surgery. Ann Thorac Surg 2020. https://doi.org/10.1016/j.athoracsur.2020.02.025.

Sutter M, Eggspuehler A, Jeszenszky D, Kleinstueck F, Fekete TF, Haschtmann D, et al. The impact and value of uni- and multimodal intraoperative neurophysiological monitoring (IONM) on neurological complications during spine surgery: a prospective study of 2728 patients. Eur Spine J 2019;28:599–610. https://doi.org/10.1007/s00586-018-5861-0.

Szelényi A, Langer D, Kothbauer K, De Camargo AB, Flamm ES, Deletis V. Monitoring of muscle motor evoked potentials during cerebral aneurysm surgery: intraoperative changes and postoperative outcome. J Neurosurg 2006;105:675–81. https://doi.org/10.3171/jns.2006.105.5.675.

Takahashi M, Imagama S, Kobayashi K, Yamada K, Yoshida G, Yamamoto N, et al. Validity of the Alarm Point in Intraoperative Neurophysiological Monitoring of the Spinal Cord by the Monitoring Working Group of the Japanese Society for Spine Surgery and Related Research. Spine (Phila Pa 1976) 2021;46:E1069–76. https://doi.org/10.1097/BRS.0000000000004065.

Thiagarajan K, Cheng HL, Huang JE, Natarajan P, Crammond DJ, Balzer JR, et al. Is two really better than one? Examining the superiority of dual modality neurophysiological monitoring during carotid endarterectomy: A meta-analysis. World Neurosurg 2015;84. https://doi.org/10.1016/j.wneu.2015.08.040.

Thirumala PD, Ahmad AI, Roy PP, Balzer JR, Crammond DJ, Anetakis KM, et al. Predictive Value of Multimodality Intraoperative Neurophysiological Monitoring During Cardiac Surgery. J Clin Neurophysiol 2021. https://doi.org/10.1097/WNP.0000000000000875.

Thirumala Parthasarathy D., Carnovale G, Habeych ME, Crammond DJ, Balzer JR. Diagnostic accuracy of brainstem auditory evoked potentials during microvascular decompression. Neurology 2014;83:1747–52. https://doi.org/10.1212/WNL.0000000000000961.

Thirumala P.D., Carnovale G, Habeych ME, Crammond DJ, Balzer JR. Diagnostic accuracy of brainstem auditory evoked potentials during microvascular decompression. Neurology 2014;83. https://doi.org/10.1212/WNL.0000000000000961.

Thirumala PD, Melachuri SR, Kaur J, Ninaci D, Melachuri MK, Habeych ME, et al. Diagnostic Accuracy of Somatosensory Evoked Potentials in Evaluating New Neurological Deficits After Posterior Cervical Fusions. Spine (Phila Pa 1976) 2017;42:490–6. https://doi.org/10.1097/BRS.0000000000001882.

Thirumala PD, Natarajan P, Thiagarajan K, Crammond DJ, Habeych ME, Chaer RA, et al. Diagnostic accuracy of somatosensory evoked potential and electroencephalography during carotid endarterectomy. Neurol Res 2016;38. https://doi.org/10.1080/01616412.2016.1200707.

Wang S, Yang Y, Li Q, Zhu J, Shen J, Tian Y, et al. High-Risk Surgical Maneuvers for Impending True-Positive Intraoperative Neurologic Monitoring Alerts: Experience in 3139 Consecutive Spine Surgeries. World Neurosurg 2018;115:e738–47. https://doi.org/10.1016/j.wneu.2018.04.162.

Wi SM, Lee H-J, Kang T, Chang SY, Kim S-M, Chang B-S, et al. Clinical Significance of Improved Intraoperative Neurophysiological Monitoring Signal during Spine Surgery: A Retrospective Study of a Single-Institution Prospective Cohort. Asian Spine J 2020;14:79–87. https://doi.org/10.31616/asj.2019.0025.

Wiedemayer H, Fauser B, Sandalcioglu IE, Schäfer H, Stolke D. The impact of neurophysiological intraoperative monitoring on surgical decisions: A critical analysis of 423 cases. J Neurosurg 2002;96:255–62. https://doi.org/10.3171/jns.2002.96.2.0255.

Wilent WB, Tesdahl EA, Harrop JS, Welch WC, Cannestra AF, Poelstra KA, et al. Utility of motor evoked potentials to diagnose and reduce lower extremity motor nerve root injuries during 4,386 extradural posterior lumbosacral spine procedures. Spine J 2020. https://doi.org/10.1016/j.spinee.2019.08.013.

Xiang B, Jiao S, Zhang Y, Wang L, Yao Y, Yuan F, et al. Effects of desflurane and sevoflurane on somatosensory-evoked and motor-evoked potential monitoring during neurosurgery: a randomized controlled trial. BMC Anesthesiol 2021;21:240. https://doi.org/10.1186/s12871-021-01463-x.
