## Supplementary Table 2 for "Standards for Reporting Diagnostic Accuracy Studies Using Intraoperative Neurophysiological Monitoring"

**Supplementary Table 2. Anesthesia reporting elements for STARD‑IONM**

| **Domain** | **Minimum** | **Enhanced** | **Ideal prospective** |
| --- | --- | --- | --- |
| Baseline timing | Report when baselines were obtained and whether they were reset. | Report timing relative to induction, stabilization, positioning, and incision, and note modality-specific reacquisition after major anesthetic, physiological, or positioning changes. | Provide time-stamped synchronization of baseline acquisition, baseline resets, and subsequent epochs with anesthetic and surgical events. |
| Anesthetic technique | State the technique (TIVA, volatile, or mixed). | Justify deviations from the planned technique. | Report protocol adherence and reasons for protocol modifications in real time. |
| TIVA agents | State hypnotic and opioid used, if applicable. | Specify delivery method, model/mode, targets, total dose, bolus policy, and major intraoperative changes. | Provide time-stamped infusion and bolus logs synchronized with IONM epochs and, if available, effect-site concentration estimates. |
| Adjuvants | Report whether adjuvants were used, if applicable. | Specify agent(s), dose, and infusion rate. | Report timing of initiation, dose adjustments, and relation to IONM changes. |
| Volatile anesthesia | State volatile agent and whether nitrous oxide was used. | Report mean end-tidal concentration and age-adjusted MAC. | Provide time-stamped end-tidal concentration data synchronized with IONM epochs. |
| Neuromuscular blockade (NMB) | Report NMB agent(s) and whether blockade was present at MEP acquisition. | Specify dosing, monitoring method, and target level before/at MEP acquisition. | Provide serial quantitative monitoring values and time-stamped changes relative to IONM events. |
| Physiology – blood pressure (BP) | Report blood pressure monitoring or management relevant to IONM interpretation, if documented. | Report BP monitoring method, target ranges, and documented BP values or vasopressor use relevant to IONM interpretation. | Provide continuous time-synchronized BP waveform or interval data linked to IONM events. |
| Physiology – oxygenation & ventilation | Report oxygenation or ventilation management relevant to IONM interpretation, if documented. | Specify SpO₂ or PaO₂ targets and EtCO₂ or PaCO₂ targets. | Provide time-stamped oxygenation and ventilation data linked to IONM epochs and alerts. |
| Physiology – temperature | Report temperature monitoring or management, if documented. | State measurement site/method and active warming measures. | Provide continuous temperature data synchronized with IONM epochs. |
| Duration & timing | Report procedural timing relevant to IONM interpretation, if noted. | Report timing of major intraoperative events relevant to interpretation. | Provide a complete time-aligned intraoperative timeline linking anesthesia, physiology, IONM, and surgical events. |
| Communication & actions | Summarize documented actions taken in response to IONM changes, if noted. | Describe action type (anesthetic adjustment, BP augmentation, warming, repositioning) and timing relative to the alert. | Provide structured time-stamped communication and intervention logs synchronized with IONM changes. |

Abbreviations: TIVA, total intravenous anesthesia; MAC, minimum alveolar concentration; NMB, neuromuscular blockade; IONM, intraoperative neurophysiological monitoring; SpO₂, peripheral oxygen saturation; PaO₂, arterial oxygen tension; EtCO₂, end-tidal carbon dioxide; PaCO₂, arterial carbon dioxide tension.

Note: Centers should report actual targets, ranges, and protocols used. Age-adjusted MAC should be reported for volatile agents.
