## Supplementary Table 3 Publications for ITEM evaluation for "Standards for Reporting Diagnostic Accuracy Studies Using Intraoperative Neurophysiological Monitoring"

**Supplementary Table 3. Publications selected for STARD-2015 item evaluation**

| **Study** | **Title** | **Journal** | **DOI** |
| --- | --- | --- | --- |
| Qiu et al., 2008 | Incidence and risk factors of neurological deficits of surgical correction for scoliosis: analysis of 1373 cases at one Chinese institution | *Spine* | 10.1097/BRS.0b013e3181657d93 |
| Jo et al., 2011 | The patterns and risk factors of hearing loss following microvascular decompression for hemifacial spasm | *Acta Neurochir* | 10.1007/s00701-010-0935-8 |
| Pulli et al., 2002 | Carotid endarterectomy with contralateral carotid artery occlusion: is this a higher risk subgroup? | *Eur J Vasc Endovasc Surg* | 10.1053/ejvs.2002.1612 |
| Raynor et al., 2007 | Correlation between low triggered electromyographic thresholds and lumbar pedicle screw malposition: analysis of 4857 screws | *Spine* | 10.1097/BRS.0b013e31815a524f |
| Mattogno et al., 2023 | Reliability of intraoperative visual evoked potentials (iVEPs) in monitoring visual function during endoscopic transsphenoidal surgery | *Acta Neurochir* | 10.1007/s00701-023-05778-1 |
| Neuloh et al., 2007b | Motor tract monitoring during insular glioma surgery | *J Neurosurg* | 10.3171/jns.2007.106.4.582 |
| Romstöck et al., 2000 | Continuous electromyography monitoring of motor cranial nerves during cerebellopontine angle surgery | *J Neurosurg* | 10.3171/jns.2000.93.4.0586 |
| Wilent et al., 2020 | Utility of motor evoked potentials to diagnose and reduce lower extremity motor nerve root injuries during 4,386 extradural posterior lumbosacral spine procedures | *Spine J* | 10.1016/j.spinee.2019.08.013 |
| Blume et al., 1986 | Significance of EEG changes at carotid endarterectomy | *Stroke* | 10.1161/01.STR.17.5.891 |
| Saito et al., 2014 | Intraoperative cortico-cortical evoked potentials for the evaluation of language function during brain tumor resection: initial experience with 13 cases | *J Neurosurg* | 10.3171/2014.4.JNS131195 |
| Iorio et al., 2023 | Utility of intraoperative neurophysiological monitoring in detecting motor and sensory nerve injuries in pediatric high-grade spondylolisthesis | *Spine J* | 10.1016/j.spinee.2023.08.002 |
| Silverstein et al., 2024 | The diagnostic accuracy of neuromonitoring for detecting postoperative bowel and bladder dysfunction in spinal oncology surgery: a case series | *J Neurooncol* | 10.1007/s11060-024-04742-y |

*Abbreviations: EEG, electroencephalography; EMG, electromyography; iVEPs, intraoperative visual evoked potentials; MEP, motor evoked potential; STARD, Standards for Reporting Diagnostic Accuracy Studies.*
