## Supplementary Table 4 for "Standards for Reporting Diagnostic Accuracy Studies Using Intraoperative Neurophysiological Monitoring"

**Supplementary Table 4. Domain-specific reference-standard examples for IONM diagnostic-accuracy studies.**

| **Surgical domain** | **Reference standard and assessment method** | **Assessor and timing** | **Positivity / severity** | **Duration / permanence** |
| --- | --- | --- | --- | --- |
| Spine / spinal cord | New postoperative neurologic deficit. Prospective standardized neurologic examination comparing motor and sensory function to preoperative baseline, with 3-month follow-up to classify deficits as transient or permanent.(Sutter et al., 2019) | Spine surgeons and neurologists. Immediate postoperative, discharge, and 3-month follow-up. | New or worsened motor or sensory deficit versus preoperative baseline; permanent deficit defined as persisting at 3 months. | Transient (resolved within 3 months) or permanent (persisting at 3 months). |
| Pedicle screw studies | Radiographic malposition and/or neurologic deficit. Postoperative CT for pedicle breach, complemented by neurologic examination for radicular pain, weakness, or sensory loss(Yongjun et al., 2023) | Spine surgeons for CT evaluation and clinical examination. Early postoperative CT and follow-up neurologic assessment. | Prespecified cortical breach definition; primary reference standard may be radiographic malposition, symptomatic malposition, or both. | Transient symptoms, persistent symptoms, need for revision, or persistent deficit at follow-up. |
| Cranial nerve / skull base | Postoperative cranial nerve dysfunction. Structured cranial nerve examination using a validated severity scale (e.g., House–Brackmann for facial nerve)(Greve et al., 2021) | Surgeon, neuro-otologist, or neurologist. Early postoperative assessment and follow-up (e.g., day 90) to distinguish transient from persistent dysfunction. | Any worsening versus clinically meaningful worsening (e.g., HB ≥IV with incomplete eye closure); severity grades reported when available. | Transient, improving, or persistent cranial nerve dysfunction on follow-up. |
| Brain tumor / awake language mapping | New or worsened postoperative language impairment. Formal pre- and postoperative language assessment comparing language function before and after surgery (Seidel et al., 2024) | Neuropsychologist, speech-language pathologist, or neurosurgeon trained in language assessment. Early postoperative and follow-up assessment. | New or worsened aphasia on formal postoperative language assessment compared with preoperative baseline; severity graded by standardized testing when available. | Transient, improving, or persistent language deficit trajectory. |
| Vascular surgery | Perioperative stroke or TIA. Postoperative neurologic assessment in patients with neurologic complaints or overt signs after CEA; stroke confirmed by clinical evaluation (Moehl et al., 2022) | Neurology and vascular surgery team at the treating institution. Immediate postoperative period; TIA defined as deficit resolving within 24 hours; perioperative stroke assessed within 30 days. | Any new focal neurologic deficit of abrupt onset after CEA; TIA distinguished from stroke by duration (<24 hours vs ≥24 hours). | TIA (resolved <24 hours) or persistent stroke. |
| Peripheral or cranial nerves | Postoperative vocal fold paresis. Postoperative laryngoscopy to evaluate vocal fold mobility after recurrent laryngeal nerve monitoring during thyroid surgery (Stopa & Barczyński, 2017) | Otolaryngologist or surgeon trained in laryngoscopy. Postoperative laryngoscopy in early postoperative period and at follow-up. | Vocal fold paresis or paralysis on laryngoscopy; IONM loss of signal defined as V2 amplitude ≤100 μV. | Transient (resolved at follow-up) or permanent vocal fold paresis. |

*Reference standards should be selected for their relevance to patient-centered outcomes, including motor and sensory function, communication ability, voice and swallowing function, and independence in daily activities. Examples are illustrative and not exhaustive; authors should justify their choice of reference standard in the context of the specific IONM modality and surgical procedure under study.*

*Abbreviations: CEA, carotid endarterectomy; CT, computed tomography; HB, House–Brackmann; IONM, intraoperative neurophysiological monitoring; TIA, transient ischemic attack.*

Greve, T., Wang, L., Katzendobler, S., Geyer, L. L., Schichor, C., Tonn, J. C., & Szelényi, A. (2021). Bilateral and Optimistic Warning Paradigms Improve the Predictive Power of Intraoperative Facial Motor Evoked Potentials during Vestibular Schwannoma Surgery. *Cancers*, *13*(24), 6196. https://doi.org/10.3390/cancers13246196

Moehl, K., Shandal, V., Anetakis, K., Paras, S., Mina, A., Crammond, D., Balzer, J., & Thirumala, P. D. (2022). Predicting transient ischemic attack after carotid endarterectomy: The role of intraoperative neurophysiological monitoring. *Clinical Neurophysiology*, *141*, 1–8. https://doi.org/10.1016/j.clinph.2022.06.010

Seidel, K., Wermelinger, J., Alvarez-Abut, P., Deletis, V., Raabe, A., Zhang, D., & Schucht, P. (2024). Cortico-cortical evoked potentials of language tracts in minimally invasive glioma surgery guided by Penfield stimulation. *Clinical Neurophysiology*, *161*, 256–267. https://doi.org/10.1016/j.clinph.2023.12.136

Stopa, M., & Barczyński, M. (2017). Prognostic value of intraoperative neural monitoring of the recurrent laryngeal nerve in thyroid surgery. *Langenbeck’s Archives of Surgery*, *402*(6), 957–964. https://doi.org/10.1007/s00423-016-1441-0

Sutter, M., Eggspuehler, A., Jeszenszky, D., Kleinstueck, F., Fekete, T. F., Haschtmann, D., Porchet, F., & Dvorak, J. (2019). The impact and value of uni- and multimodal intraoperative neurophysiological monitoring (IONM) on neurological complications during spine surgery: a prospective study of 2728 patients. *European Spine Journal : Official Publication of the European Spine Society, the European Spinal Deformity Society, and the European Section of the Cervical Spine Research Society*, *28*(3), 599–610. https://doi.org/10.1007/s00586-018-5861-0

Yongjun, T., Yuntian, Z., Biao, C., & Zenghui, J. (2023). Intraoperative triggered electromyographic monitoring of pedicle screw efficiently reduces the lumbar pedicle breach and re-operative rate-a retrospective analysis based on postoperative computed tomography scan. *BMC Musculoskeletal Disorders*, *24*(1), 535. https://doi.org/10.1186/s12891-023-06658-6
